# Evidence of Protective Role of Ultraviolet-B (UVB) Radiation in Reducing COVID-19 Deaths

**DOI:** 10.1101/2020.05.06.20093419

**Authors:** Rahul Kalippurayil Moozhipurath, Lennart Kraft, Bernd Skiera

**Author notes:** Theodor-W.-Adorno-Platz 4, 60629 Frankfurt, Germany.

## Abstract

**Background:** Research is ongoing to identify an effective way to prevent or treat COVID-19, but thus far these efforts have not yet identified a possible solution. Prior studies indicate the protective role of Ultraviolet-B (UVB) radiation in human health, mediated by vitamin D synthesis. In this study, we empirically outline a negative association of UVB radiation as measured by ultraviolet index (UVI) with the number of deaths attributed to COVID-19 (COVID- 19 deaths).

**Methods:** We carry out an observational study, applying a fixed-effect log-linear regression model to a panel dataset of 152 countries over a period of 108 days (n=6524). We use the cumulative number of COVID-19 deaths and case-fatality rate (CFR) as the main dependent variables to test our hypothesis and isolate UVI effect from potential confounding factors such as underlying time trends, country-specific time-constant and time-varying factors such as weather.

**Findings:** After controlling for time-constant and time-varying factors, we find that a permanent unit increase in UVI is associated with a 1.2 percentage points decline in daily growth rates of cumulative COVID-19 deaths [p < 0.01] as well as a 1.0 percentage points decline in the daily growth rates of CFR [p < 0.05]. These results represent a significant percentage reduction in terms of the daily growth rates of cumulative COVID-19 deaths (−11.88%) and CFR (−38.46%). Our results are consistent across different model specifications.

**Interpretation:** We find a significant negative association between UVI and COVID-19 deaths, indicating evidence of the protective role of UVB in mitigating COVID-19 deaths. If confirmed via clinical studies, then the possibility of mitigating COVID-19 deaths via sensible sunlight exposure or vitamin D intervention will be very attractive because it is cost-effective and widely available.

## 1 Introduction

COVID-19 is causing significant economic, healthcare and social disruption globally. However, it is not yet known how to prevent or treat COVID-19. Prior studies indicate the protective role of Ultraviolet-B (UVB) radiation in human health. UVB radiation exposure is a major source of vitamin D, which increases immunity and reduces the likelihood of severe infections and mortality.

A recent COVID-19 study indicates abnormally high case-fatality-rate (CFR) of 33.7% among nursing home residents ^1^, which is consistent with the studies indicating higher prevalence of vitamin D deficiency among them, due to lower mobility ^2,3^ Increasingly, studies establish a link between vitamin D deficiency and comorbidities such as cardiovascular disease ^4^, hypertension ^5^, obesity ^2,6^, type 1, and type 2 diabetes ^7^ This evidence is consistent with the clinical studies in China and Italy that indicate comorbidities such as hypertension, diabetes and cardiovascular diseases could be important risk factors for critical COVID-19 cases ^8–10^. Epidemiology of COVID-19 provides evidence that vitamin D might be helpful in reducing risk associated with COVID-19 deaths^11,12^. If such a link is true, then it will be cost-effective to mitigate COVID-19 via sensible exposure to sunlight or via vitamin D nutritional intervention. Yet, to the best of our knowledge, so far, no empirical study has used data across many countries to explore the association between UVB radiation as measured by ultraviolet index (UVI) and the number of deaths attributed to COVID-19 (COVID-19 deaths).

The aim of this study is therefore to examine the relation of UVB radiation, as measured by ultraviolet index (UVI), with the number of COVID-19-deaths. The results of our study demonstrate that a one-unit increase in UVI is associated with a 1.2 percentage points decline in daily growth rates of cumulative COVID-19 deaths. The robustness checks show similar effect of UVI on case fatality rate (effect size: -0.010) and the results are consistent across a variety of different model specifications (effect size: -0.006 to -0.012).

A major threat to identifying the effect of UVB with the number of COVID-19 deaths is the presence of time trends, which could affect UVI as well as the number of COVID-19 deaths. For example, many countries affected by COVID-19 are in the northern hemisphere leading to a natural phenomenon that UVI increases over time. In addition, growth rates of the cumulated COVID-19 deaths are decreasing over time. This negative correlation between UVI and the cumulated COVID-19 deaths due to time is the source of the identification problem. We address this problem through our statistical analysis in which we flexibly isolate UVI from linear or non-linear time trends which can be either similar across countries or even country-specific.

## 2 Importance of UVB Radiation for Human Health

Prior studies find that UVB radiation plays a protective role in human health because it reduces the severity of immune diseases ^13^, reduces the risk of getting cancer - e.g., prostate cancer ^14^ and dying from cancer ^15,16^ and may reduce the prevalence of hypertension^17^.

Humans receive vitamin D either from their diet (natural food, fortified food or supplements) or from skin synthesis by solar UVB radiation exposure ^18^. In general, skin synthesis is the major source of vitamin D ^19,20^, as the dietary intake is usually insufficient ^21^. Various studies consider that UVB exposure twice a week is sufficient to maintain vitamin D levels ^21^ and vitamin D once produced can be stored in body fat and can be utilized later^21^, indicating a lagged effect of UVB.

UVB radiation shows significant variation according to latitude, seasons and time of the day. Specifically, during winter months in northern latitudes (e.g., above 35° latitude - Oklahoma - US), the ozone absorbs most of the UVB^22^, leading to reduced likelihood of UVB radiation exposure and thereby insufficient vitamin D synthesis as indicated in Figure 1.

**Figure 1:**
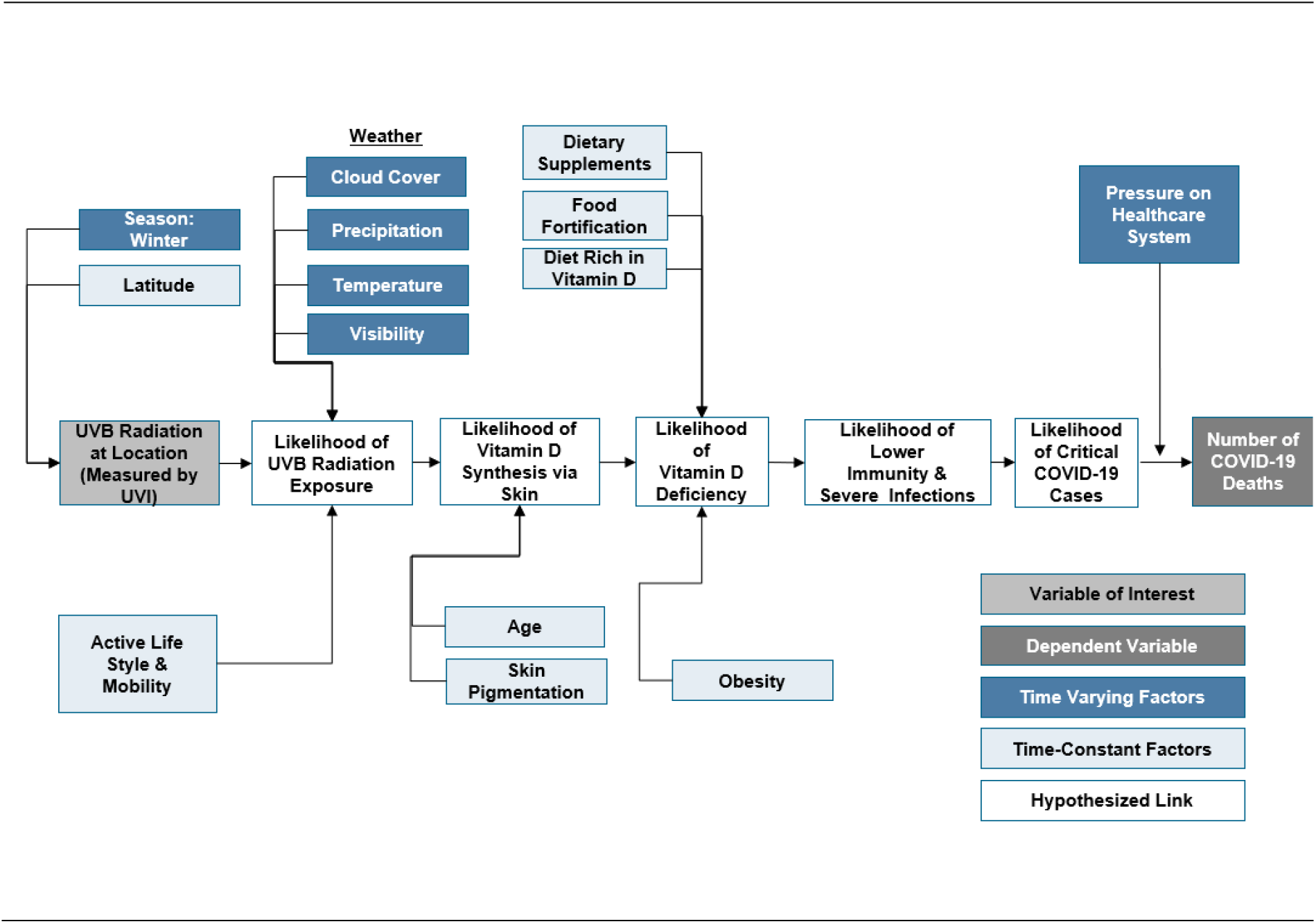
Explanation of Protective Role of Ultraviolet-B (UVB) Radiation in COVID-19 Deaths Mediated by Vitamin D Synthesis & Deficiency

Other weather factors such as cloud cover, precipitation, visibility and temperature influence the likelihood of exposure to UVB radiation and thereby vitamin D deficiency due to reduced skin synthesis. For example, clouds not only reduce the amount of UVB radiation but also the likelihood of UVB radiation exposure as people are more likely to undertake outdoor activities on less cloudy days. Lifestyle and mobility also influences the likelihood of UVB radiation exposure ^3,23,24^ Similarly, the likelihood of vitamin D deficiency also increases with age ^21^, skin pigmentation ^25^ and obesity due to reduced skin synthesis ^26^.

In Figure 1, we summarize these different factors explaining the potential protective role of UVB radiation in reducing COVID-19 deaths, mediated by vitamin D synthesis and deficiency. Since UVB radiation exposure is a major source of vitamin D, an increase in the likelihood of skin exposure to UVB radiation increases vitamin D synthesis, thereby reducing the likelihood of vitamin D deficiency. Therefore, different time varying and time-constant factors influencing the UVB radiation variation and exposure also influence the likelihood of vitamin D synthesis and thereby deficiency. Prior studies indicate that vitamin D deficiency increases the likelihood of weakened immune response ^18,27,28^, infectious diseases in the upper respiratory tract ^21,29,30^ and the severity as well as mortality in critically ill patients ^31^. Therefore, we expect that an increased skin synthesis of vitamin D due to increased UVB radiation increases the likelihood of immunity and reduces the likelihood of severe infections, thereby reducing the critical COVID-19 cases. Thus, we anticipate that an increase in UVB radiation as measured by ultraviolet index (UVI) relates to a reduction of the number of COVID-19 deaths.

## 3 Methods

### 3.1 Description of Data

In order to identify the relation of UVB radiation and COVID-19 deaths, we constructed the dataset outlined in Table 1. We collected data covering 108 days from 22 January 2020 until 8 May 2020 across 183 countries of which 158 reported COVID-19 deaths prior to 8 May 2020 and of which 152 reported more than 20 COVID-19 infections prior to 8 May 2020. We focus on those 152 countries to ensure that the results are not biased by countries that are at a very early stage of COVID-19 outbreak, which would limit data points with respect to COVID-19 deaths. In addition, we drop the first 20 daily observations of every country after that country reported the first COVID-19 infection to further ensure that results are not biased by the observations at the very early stage of the COVID-19 outbreak.

**Table 1:** Summary of Dataset

|  |  |
| --- | --- |
| Number of countries in the world | 195 |
| Number of countries in our dataset | 183 |
| ... > 0 cumulated COVID-19 deaths before 8 May 2020 | 158 |
| ... > 20 cumulated COVID-19 infections before 8 May 2020 | 152 |
| Covered time-period | 22 January 2020 - 8 May 2020<br>(108 days) |
| Granularity of data | Daily |
| COVID-19 data source | https://github.com/CSSEGISandData/COVID-19 |
| Weather data source | https://darksky.net/ |

The corresponding country level data consist of the cumulative daily COVID-19 deaths and infections, the daily ultraviolet index (UVI), which is closely connected to the daily UVB radiation, and a set of control variables such as daily weather parameters such as precipitation index, cloud index, ozone level, visibility level, humidity level, as well as minimum and maximum temperature.

We present descriptive statistics of the dataset in Table 2. As of 8^th^ of May, the cumulative COVID-19 deaths of these 152 countries were on average 1,808 and the growth rate of COVID-19 deaths on May 8 was on average 2.6% as compared to the average growth rate of COVID-19 deaths across countries and time which was 10.1%. The cumulative COVID-19 infections were on average 25.888. The case-fatality-rate (CFR), as measured by the cumulative COVID-19 deaths divided by the cumulative COVID-19 infections per country, was on average 4.3%. The growth rate of CFR on May 8 was on average -1.1% as compared to the growth rate of CFR across countries and time which was 2.6%. We use cumulative COVID-19 deaths as the main dependent variable to test our hypothesis linking UVB radiation to COVID-19 deaths and use the CFR to test the consistency of our results. On average, the first reported COVID-19 infection in each country happened 68.15 days before 8 May 2020. UVI is on average 6.81 representing a moderate to high risk of harm from unprotected sun exposure.

**Table 2:** Descriptive Statistics of Dataset

| Variable | Number of Countries | Number of Observations | Mean | Std. Dev. | Min | Max |
| --- | --- | --- | --- | --- | --- | --- |
| Cumulated COVID-19 deaths on 8 May | 152 | 152 | 1,808 | 7,779 | 1 | 77,180 |
| Growth rate of cumulative COVID-19 deaths on 8 May | 152 | 152 | 0.026 | 0.051 | 0 | 0.4 |
| Daily growth rate of cumulative COVID-19 deaths | 152 | 6,589 | 0.101 | 0.259 | -1 | 9 |
| Cumulated COVID-19 Infections on 8 May | 152 | 152 | 25,888 | 111,303 | 23 | 1,283,929 |
| Daily CFR on 8 May | 152 | 152 | 0.043 | 0.037 | 0.001 | 0.206 |
| Growth rate of CFR on 8 May | 152 | 152 | -0.011 | 0.057 | -0.421 | 0.238 |
| Daily growth rate of CFR | 152 | 6,589 | 0.026 | 0.200 | -1 | 5.935 |
| Time-passed by from first reported infection until 8 May | 152 | 152 | 68.15 | 17.59 | 29 | 108 |
| Daily Ultraviolet Index (UVI) | 152 | 7,471 | 6.81 | 3.05 | 0 | 14 |
| Daily precipitation index | 152 | 7,471 | 0.29 | 0.31 | 0 | 1 |
| Daily cloud index | 152 | 7,471 | 0.50 | 0.30 | 0 | 1 |
| Daily ozone level | 152 | 7,471 | 308.48 | 47.06 | 235.5 | 472.7 |
| Daily Visibility level | 152 | 7,471 | 15.29 | 2.17 | 0.117 | 16.09 |
| Daily humidity level | 152 | 7,471 | 0.63 | 0.20 | 0.04 | 1 |
| Minimum temperature per day within a country | 152 | 7,471 | 12.45 | 9.78 | -23.45 | 30.87 |
| Maximum temperature per day within a country | 152 | 7,471 | 22.83 | 10.47 | -16.41 | 45.82 |

### 3.2 Illustration of Ultraviolet Index (UVI) and COVID-19 Deaths

The graph on the top of Figure 2 shows the cumulative COVID-19 deaths and the associated daily growth rates for Italy from 26 February 2020 until 8 May 2020. As time progresses, the cumulative COVID-19 deaths increase but at a slower rate. Initially, the growth rate is high at 41.67% (growth rate from 26 February to 27 February) and it gradually slows to 0.81% (growth rate from 7 April to 8 May).

**Figure 2:**
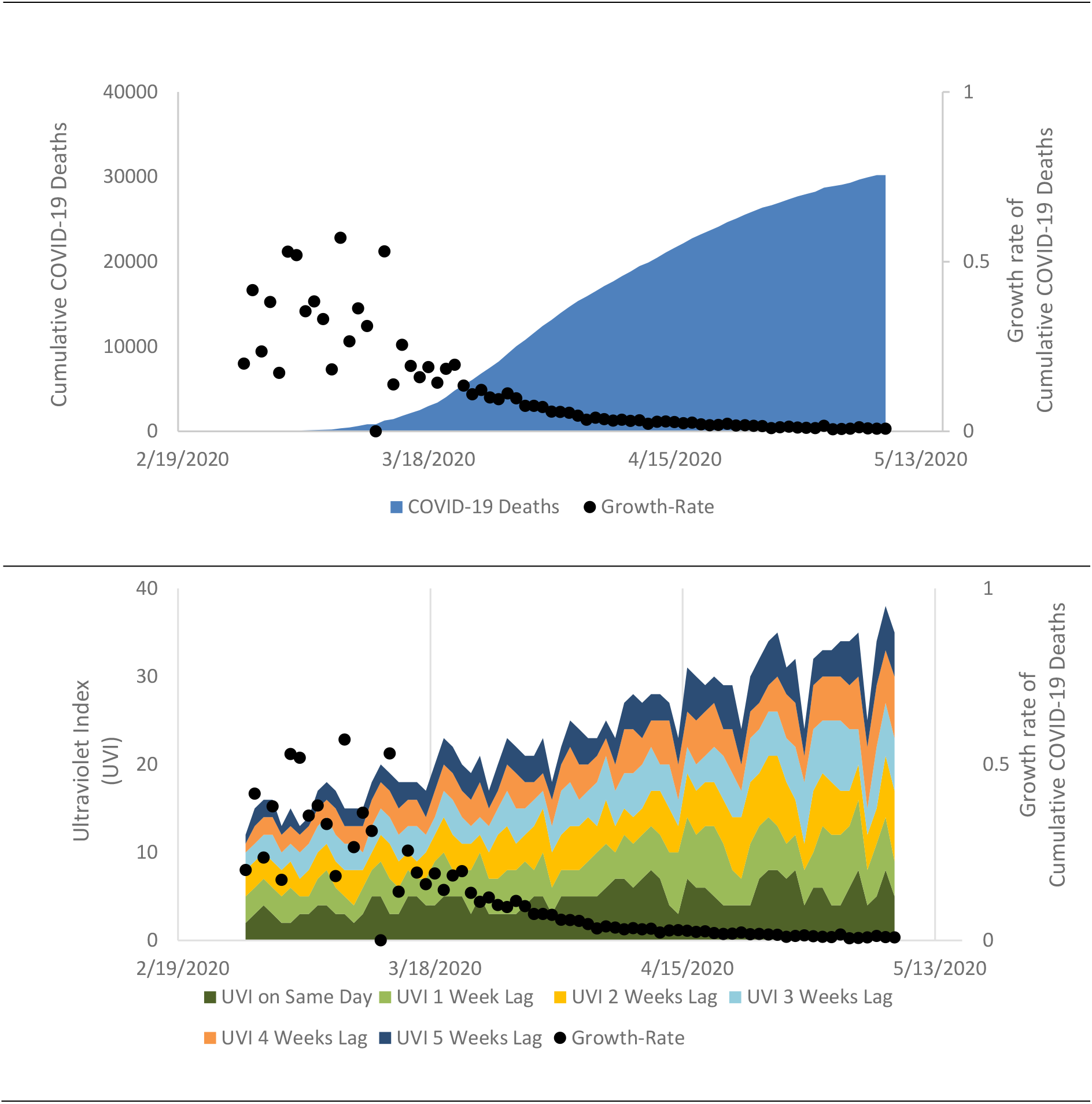
Number of COVID-19 deaths, Growth Rates and ultraviolet index (UVI) for Italy

The graph on the bottom of Figure 2 shows the daily growth rates and daily UVI for Italy as well as the UVI values lagged by one, two, three, four and five weeks respectively. It is important to consider the lagged effect of UVI because synthesized vitamin D is cumulative and can be stored in body fat to be used later^21^. Therefore, it seems more plausible that an increase of UVI today will continue to support an individual’s immunity later i.e., two-or more weeks’ time. Furthermore, the likelihood of skin synthesis is low in severely infected people, while they are hospitalized, indicating the importance of lagged UVI values.

It is evident that the growth rates slow down over time, as counter-measures imposed by governments take effect, which results in lower infection rates and lower mortality rates. At the same time, the UVI is increasing due to seasonal changes in the northern hemisphere countries. In order to approximate the association of UVI, we need to isolate it from the underlying time-trends, which are potentially affecting both UVI as well as the growth rates of cumulative COVID-19 deaths.

## 4 Results

We estimate the effect of UVI on the cumulative COVID-19 deaths by using log-linear fixed-effects regression. The effect of UVI is isolated from time-constant country-specific factors (see Figure 1) by using a within-transformation of the transformed structural model as outlined in equation (1) in *Supplementary Appendix*. Further, we use the partialling-out property to isolate the effect of UVI from all linear as well as some non-linear effects of time varying factors such as weather and time, which may confound the results. Our statistical analysis is outlined in detail in *Description of Methodology* section in *Supplementary Appendix*.

The key finding is the significant negative long-run association of UVI on cumulative COVID-19 deaths. As we outline in the *Identification of UVI Effect* section in *Supplementary Appendix*, the estimate is likely to identify an upper bound of the relation, indicating that the association could be even stronger. Our results presented in Table 3 suggest that a permanent unit increase of UVI is associated with a decline of 1.2 percentage points in daily growth rates of cumulative COVID-19 deaths [p < 0.01]. Relative to the average daily growth rate of cumulative COVID-19 deaths (10.1%), this decline translates into a significant percentage change of -11.88% (=-1.2%/10.1%). We further find that a permanent unit increase of UVI is associated with a decline of 1.0 percentage points in the daily CFR growth rate [p < 0.05]. Compared with the average daily growth rate of CFR (2.6%), this decline translates into a significant percentage change of -38.46% (=-1.0%/2.6%).

**Table 3:**
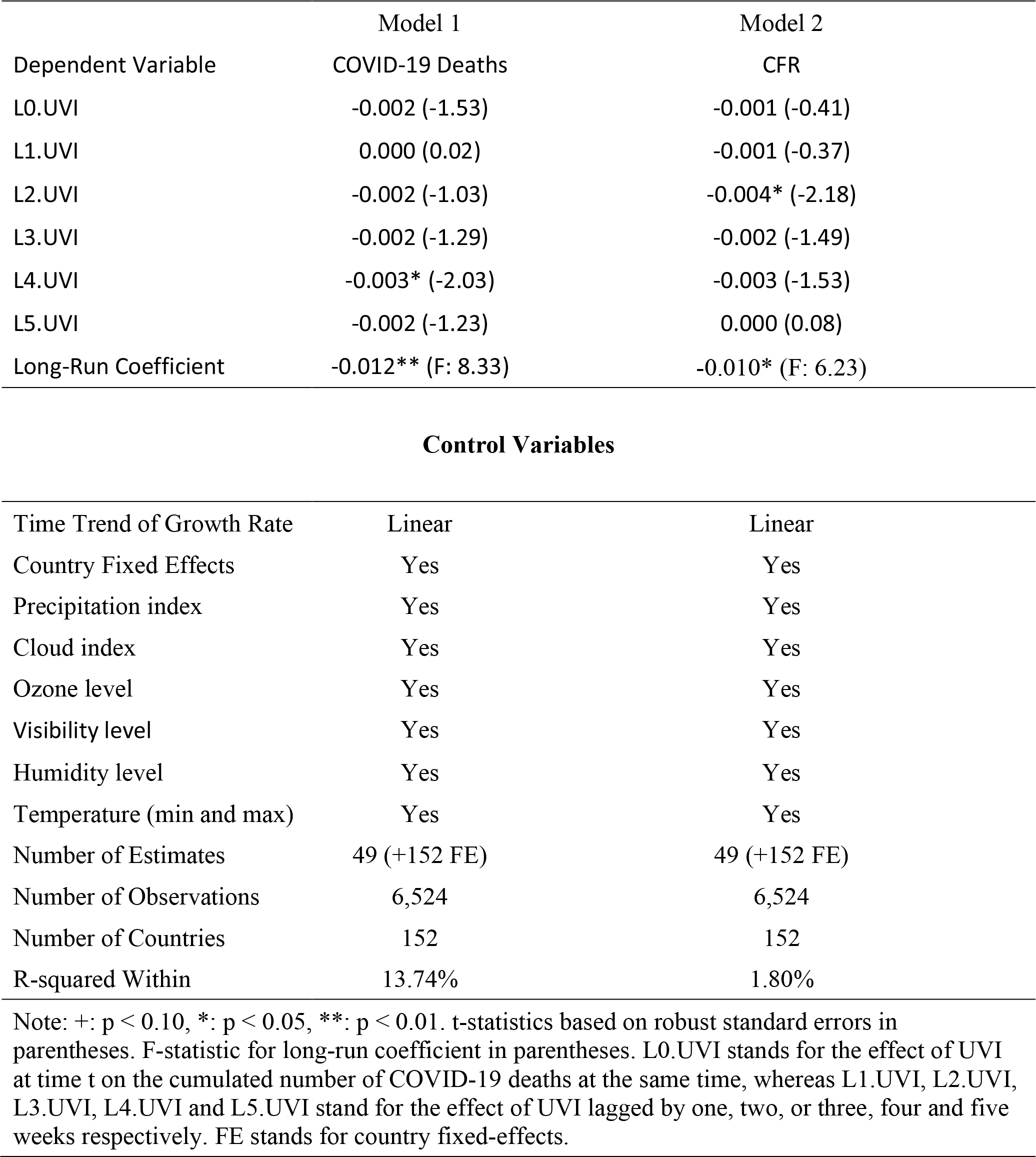
Effect of UVI on the Cumulative COVID-19 Deaths

Results indicate no significant association from an increase of UVI on cumulative COVID-19 deaths on the same day or a week ahead. This insignificant finding is consistent with the fact that severely infected people are more likely to be hospitalized and therefore less likely to be exposed to UVB radiation during their hospital stay. We further recognize that UVB radiation may not make a real difference, when someone is already severely infected and developed severe complications. The results also show that UVI has a stronger relation to COVID-19 deaths than CFR. We anticipate that the weaker association with CFR is plausible as UVI helps in vitamin D synthesis, making the infection less severe due to increased immunity, thereby prompting fewer people to take the test.

The results of the robustness checks presented in Table 2 and Table S3 (*Robustness Checks* section in *Supplementary Appendix*) suggest that the relation of UVI on cumulative COVID-19 deaths is consistent (−0.006 - −0.012) across different model specifications which isolate the association of UVI from underlying time trends in flexible ways. In fact, the most flexible model - Model 8 of Table S3 (*Robustness Checks* section in *Supplementary Appendix*) - reveals substantial and significant evidence of the UVI relation with cumulative COVID-19 deaths (−0.008, p < 0.05). A decline of 1.2 percentage points in daily growth rates of cumulative COVID-19 deaths has significant long-run effects on the cumulative COVID-19 deaths as outlined in Figure 3. In order to simulate the long-run effects, we take the average number of cumulative COVID-19 deaths across all 152 countries as of May 8, 2020, i.e., 1,808 as cumulative COVID-19 deaths at day 0 as shown in Figure 3. A scenario with a permanent unit increase of UVI over the baseline scenario of average UVI of 6.81 across countries is associated with 989 or 14.22% fewer deaths in 14 days.

**Figure 3:**
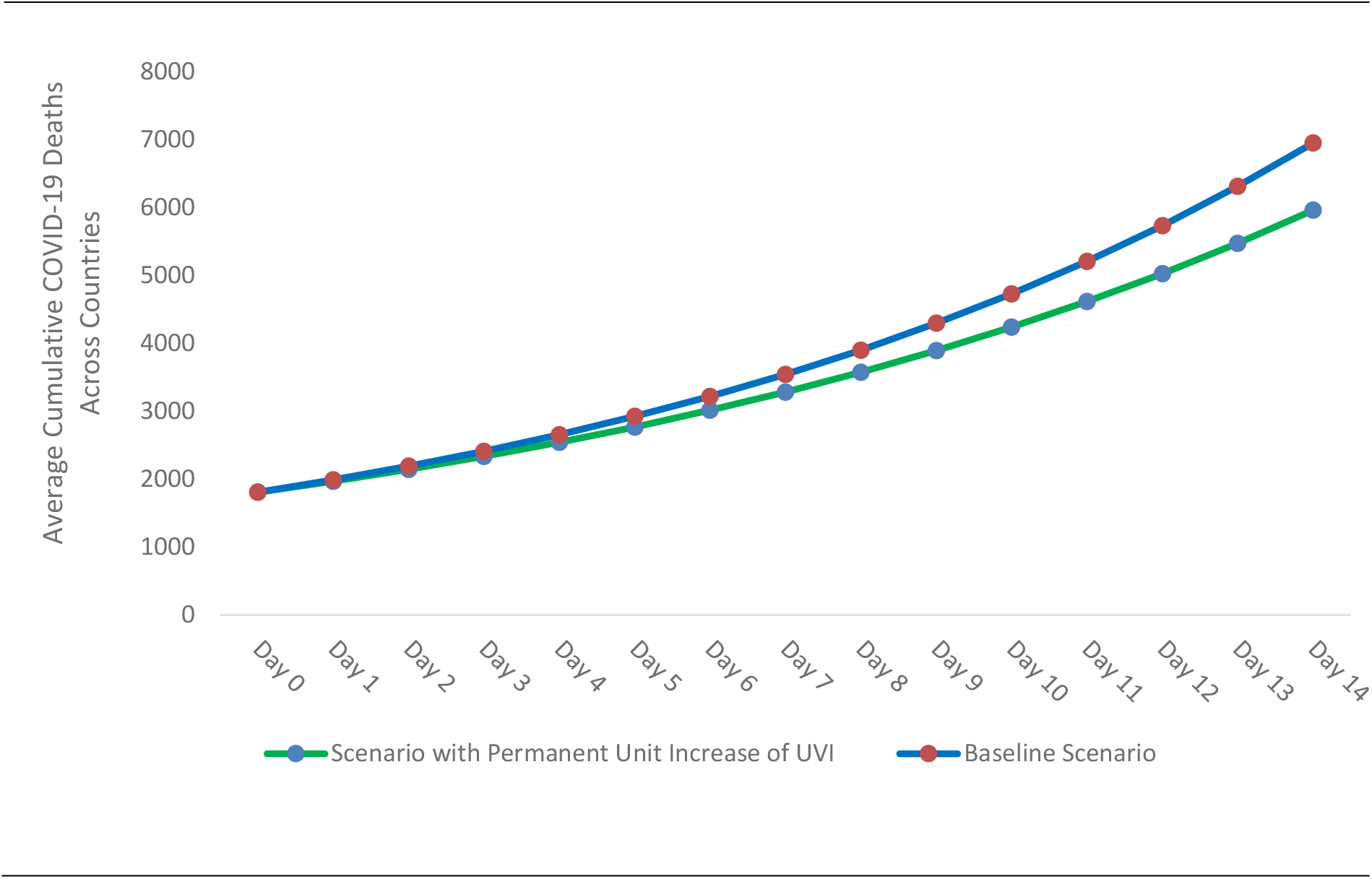
Long-run Effects of a Permanent Unit Increase of UVI on Average Cumulative COVID-19 Deaths Across Countries

## 5 Discussion

In this study, we find evidence of the protective role of UVB radiation in reducing COVID-19 deaths. Specifically, we find that a permanent unit increase in ultraviolet index (UVI) is associated with a 1.2 percentage points decline in daily growth rates of COVID-19 deaths [p < 0.01] as well as a 1.0 percentage points decline in the daily growth rates of CFR [p < 0.05]. These results translate into a significant percentage reduction in terms of the daily growth rates of cumulative COVID-19 deaths (−11.88%) and CFR (−38.46%). Our results are consistent across different model specifications.

We acknowledge that we may not be able to isolate the association of UVI with cumulative COVID-19 deaths from all confounding factors. Still, we anticipate that an increased likelihood of immunity and a reduced likelihood of infections mediated by an increased likelihood of vitamin D synthesis may plausibly explain this finding. We also acknowledge that we may not be able to rule out the possibility of mediation by other UVB induced mediators – such as cis-urocanic acid, nitric oxide ^13,32^ Therefore, further clinical studies – observational or randomized controlled trials - are required to establish the casual relationship of vitamin D deficiency and COVID-19 deaths, potentially leading to a cost-effective policy intervention for the prevention or as a therapy for COVID-19. The possibility of mitigating COVID-19 via sensible exposure to sunlight or via vitamin D intervention seem to be very attractive from a policy maker’s perspective because of its low cost and side effects.

Various countries are implementing lockdown as a preventive measure to mitigate COVID-19 impact on healthcare system. Unfortunately, confinement at home also leads to limited UVB exposure, possibly increasing the risk of COVID-19 deaths. While sensible exposure to sunlight helps in synthesizing vitamin D, disproportionate exposure may also increase the risk of sunburn and skin cancer^21^. Countries could create awareness among the population regarding the importance of sensible exposure to sunlight, whilst continuing other measures such as social distancing as well as cautioning against disproportionate exposure. If confirmed via additional clinical studies, then countries could adopt a cost-effective vitamin D intervention program – especially among vulnerable populations with increased risk of vitamin D deficiency, e.g., elderly populations living in nursing homes, people with high body mass index, dark skinned people residing in higher latitudes, people with indoor lifestyle, or vegetarians.

## Data Availability

We will make dataset used in this study available for any future research. Interested researchers can contact one of the authors via email to get access to the data.

## 7 Acknowledgements

We would like to acknowledge Sharath Mandya Krishna, and Rukhshan Ur Rehman for their immense contribution to this paper - for providing inputs and assisting with data collection, data transformation and data engineering. We thank Matthew Little for his assistance in review. We would also like to acknowledge Michael Niekamp, Magdalena Ceklarz and Daniel Gutknecht for their valuable contributions to our paper and the discussions about COVID-19.

## 8 Author Contributions

RKM conceptualized the research idea, conducted literature research, designed theoretical framework and collected the data. LK designed empirical methods and analyzed the data. RKM and LK interpreted the results and wrote the article. BS provided critical inputs, edited and revised the article.

## 10 Additional Information

Correspondence and requests for materials should be addressed to Rahul Kalippurayil Moozhipurath.

## 11 Data Sharing

The data used in the study are from publicly available sources. Data regarding COVID-19 are obtained on 9^th^ May 2020 from *COVID-19 Data Repository* by the *Center for Systems Science and Engineering* (*CSSE*) at *Johns Hopkins University* and can be accessed at https://github.com/CSSEGISandData/COVID-19. Data regarding weather is obtained from *Dark Sky* on the 9^th^ May 2020 and can be accessed at https://darksky.net/. We will make specific dataset used in this study available for any future research. Interested researchers can contact one of the authors via email to get access to the data.

## 13 Supplementary Appendix

### 13.1 Description of Methodology

We apply a fixed-effect log-linear regression model to estimate the effect of UVI on the number of COVID-19 deaths that builds upon Figure 1 in *Manuscript*. A log-linear model increases the comparability of the growth rates of COVID-19 deaths across countries because it considers percentage rather than absolute changes over time, and percentage growth rates are more comparable across countries than absolute ones. The model isolates the effect of UVI from country-specific time-constant factors. These time-constant factors consist of the countries’ latitude and diet related effects such as dietary supplements, food fortification and diets which are rich in vitamin D. They also consist of population parameters such as how active their people’s lifestyle and mobility are and the composition of the population with respect to their age, skin pigmentation or obesity rates.

Furthermore, the model allows to control for an increasing pressure on the healthcare system over time measured by the time passed by since the first reported case of COVID-19 in the specific country. Importantly, this factor would partial out any linear change of growth rates over time that is similar across countries. Therefore, the model isolates the effect of UVI from an exponential-shaped curve which is often observed in the cumulative COVID-19 deaths over time or in the growth rates of Figure 2 in *Manuscript*. The model also isolates the effect of UVI from factors which can influence UVI or individuals’ absorption of UVB such as precipitation index, cloud index, ozone level, visibility level, humidity level, and temperature.

Because an increase of UVB today plausibly affects individuals two and three weeks later we include in our model three weekly lags of UVI and of the control variables. Thus, we use the following model to explain the number of COVID-19 deaths:

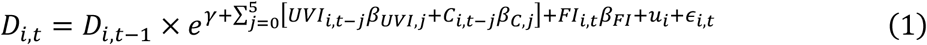

*D_i,t_* represents the cumulative COVID-19 deaths for country *i* at time point *t* (in days) and it is related to the explanatory factors via an exponential growth model on the right-hand side of the equation. The exponential growth model flexibly allows for different shapes of the cumulative COVID-19 deaths. These shapes cover the one described Figure 2 in *Manuscript* or often observed S-shaped curves which appear at later stages of COVID-19 outbreaks, depending on how flexible time is allowed to enter the exponential growth model.

The exponential growth model consists of six explanatory parts.

1. *γ* represents the daily growth rate of COVID-19 deaths from *D_i,t−_*_1_ to *D_i,t_* that is independent of the factors presented in Figure 1 in *Manuscript. γ* covers virus-specific attributes like its basic reproductive rate R_0_ combined with its lethality.
2. *UVI_i,t−j_* represents the *UVI* for a country *i* at day *t* lagged by *j* weeks. *β_UVI,j_* reflects the effect of *UVI* lagged by *j* weeks.
3. *C_i,t−j_* stands for the set of control variables. This set consists of precipitation index, cloud index, ozone level, visibility level, humidity level, as well as minimum and maximum temperature for a country *i* at day *t* lagged by *j* weeks. The vector *β_C,j_* identifies the effect of these control variables lagged by *j* weeks.
4. *FI_i,t_* stands for the time passed by since the first reported COVID-19 infection for a country *i* at day *t* and *β_FI_* identifies the associated effect.
5. *u_i_* represents time-constant country-specific factors influencing the growth rate of cumulative COVID-19 deaths (e.g., diet related effects, population parameters about their activities and demographic composition).
6. *∊_i,t_* consists of all the remaining factors that are not identified but also have an effect on the cumulative COVID-19 deaths (i.e., all non-linear differences of growth rates with respect to time and country-specific linear differences of growth rates with respect to time. They could be caused by a decreasing number of people who could potentially become infected or contagious, lockdowns in a country over time, mutation of the virus in a country over time, systematic false-reports of the dependent variable).

An appropriate transformation (see Section 13.6 for the Derivation of Equation (2)) results in the estimable equation (2).

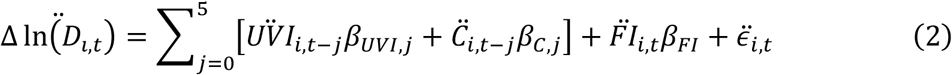

*γ* and *u_i_* do not appear in the equation anymore and a linear regression can identify all other coefficients. Equation (2) also shows why we can only use those observations where cumulative COVID-19 deaths are larger than zero. For example, the first COVID-19 death of Italy was reported on 02/21/2020. Therefore, we can only include observations of Italy starting from 02/22/2020. This condition explains the difference between the 7,471 observations of 152 countries in Table 2 and the 6,524 observations of 152 countries in Table 3 in *Manuscript*. We present an overview of how many observations of which country we use in our analysis in Table S8.

### 13.2 Derivation of Short- and Long-Run Effects

The interpretation of the coefficients of equation (2) is percentage wise and the effect of lagged variables can be separated into a short- and a long-run effect. For example, a one-unit increase of *UVI* at time s affects the cumulative COVID-19 deaths *D_t=s_* via *β_UVI_,_0_* in the short-run. After one week, the increase of *UVI* affects *D_t=s+_*_1_ firstly via *D_t=s_* and secondly via *β_UV,_*_1_ because 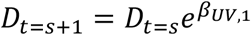 (partialling out the daily growth rate *γ* and keeping the control variables constant). Consequently, if the model consists of 3 lags, the long-run effect will be reached after 3 weeks (see Table S1). Therefore, an increase of *UVI* by one-unit increases the cumulative COVID-19 deaths approximately by (*β_UV,_*_0_ *+ β_UV,_*_1_ *+ β_UV,_*_2_ *+ β_UV,_*_3_ + *β_UV,_*_4_ + *β_UV,_*_5_) × 100% percent in the long-run (the exact number is (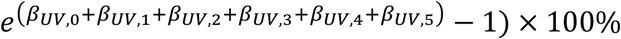). In comparison, a permanent increase of *UVI* by one-unit increases the cumulative COVID-19 deaths approximately by (*β_UV,_*_0_ *+ β_UV,_*_1_ *+ β_UV,_*_2_ *+ β_UV,_*_3_ + *β_UV,_*_4_ + *β_UV,_*_5_) × 100% each day.

### 13.3 Identification of Effect of Ultraviolet Index (UVI)

The key assumption that is required to identify a causal effect of UVI on the cumulative COVID-19 deaths is that *UVI_i,t−s_* is uncorrelated to *ϵ_i,t_* at all points in time. This means that 1) past or future unexplained parts of *D_i,t_* must not affect *UVI_i,t_*. These unexplained parts would appear in *ϵ_i,t_* and be correlated with *UVI_i,t−s_* for some *s*. The key assumption requires further, that 2) there is no factor affecting *D_i,t_* which also influences *UVI_i,t_* in addition to country-specific time-constant factors or those variables which we include in the analysis. This additional factor would appear in *ϵ_i,t_* and correlate with *UVI_i,t−s_* for some *s*. Both requirements are always satisfied, if UVI is randomly distributed after controlling for an appropriate set of control variables^1^.

The first assumption is likely satisfied because the cumulative COVID-19 deaths cannot influence UVI. However, there have been structural changes with respect to the behavior of individuals because the number of COVID-19 deaths likely influences them. Currently, individuals of regions where COVID-19 is present are less likely to go outside. However, going outside is a precondition for individuals to absorb UVB. Suppose the effect of UVI is *β_high_* high for the first ten observations and *β_low_* for the subsequent twenty observations, because individuals do not go outside and absorb UVB. Then, the estimate *β_est_* recovers the weighted average of both effects, meaning that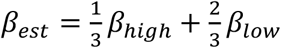, but would not be biased due to the behavioral change.

The second assumption could be violated by changes in the growth rates of the cumulative COVID-19 deaths with respect to countries over time. Such changes could be growth rates which are 1) country-specific and time-constant, 2) linearly time-varying but similar across countries, 3) non-linearly time-varying but similar across countries, and 4) linearly or nonlinearly time-varying and country-specific. Such changes in the growth rates threaten a causal identification of the effect of UVI because UVI increases over time as summer is coming closer (in the countries on the Northern hemisphere). Our main model specification isolates the effect of UVI from changes in the growth rates of type 1) and type 2) by partialling out any country-specific time-constant differences of growth rates and linear changes of growth rates with respect to time that are similar across countries. In our robustness checks we increase flexibility of the model to also capture country-specific linear differences of growth rates with respect to time as well as some non-linear differences of growth rates with respect to time, which are either similar across countries or even country-specific.

Cloud or precipitation index could also violate condition 2). On the one hand, a high cloud coverage and precipitation today decrease the future number of infections because individuals are less likely to go outside and get infected today and die in future. On the other hand, both indices decrease UVI, because they absorb the radiation. Therefore, these two relationships could create an upward bias in the estimate of UVI. Consequently, controlling for the cloud and precipitation index mitigates the bias and makes a causal identification more plausible.

The air quality could violate condition 2). The decrease in traffic over time leads to an improvement of the air quality and air quality is likely to reduce the cumulative COVID-19 deaths^2^. Because UVI increases over time, the air quality could be positively correlated with UVI. These relationships could cause an upwards bias in the estimate of UVI meaning that the true effect of UVI is lower that the estimate. Therefore, negative estimates can be considered conservative.

A country-specific mutation of the virus over time could also violate assumption 2. If a mutation increased the cumulative COVID-19 deaths, then it could positively correlate with UVI due to time. This relationship would create an upward bias. Therefore, negative estimates of the effect of UVI can be considered conservative. Another threat to our identification is a potential systematic false reporting about the cumulative COVID-19 deaths. In the beginning of the crisis it seems likely that not all deaths have been tested for COVID-19 (so that the reported number of COVID-19 deaths is smaller than the true value) while nowadays all deaths which are tested positively for COVID-19 are reported as a COVID-19 death, even though not entirely caused by it (i.e., reported COVID-19 deaths is higher than true value). This positive correlation of measurement error with time would generate an upward bias of the estimate of the effect of UVI. Therefore, negative estimates can be considered conservative.

### 13.4 Model Selection to Identify the Effect of Ultraviolet Index (UVI) on the Cumulative COVID-19 Deaths

We estimate equation (2) up to a lag of 8 weeks and decided to choose models with 5 lags and all control variables as presented in Table S2. On the one hand, we did not find major changes with respect to the size and statistical significance of the estimate of UVI. On the other hand, including more lags increases the number of estimates which decreases their accuracy such that a more parsimonious model is favorable.

### 13.5 Robustness Checks

To examine the robustness of our results we change the dependent variable into the case-fatality-rate (CFR). CFR is defined as the cumulative COVID-19 deaths divided by the cumulative COVID-19 infections. Therefore, CFR of country *i* day *t* is calculated as 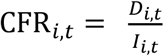, where *I_i_*,*_t_* stands for the cumulative COVID-19 infections. It is a common measure to assess the severity of diseases because a high CFR leads to a high number of cumulative COVID-19 deaths. Its advantage to the cumulative COVID-19 deaths is that it relates the cumulative number of deaths to the cumulative number of infections for a disease. Therefore, it helps to isolate the effect of UVI on cumulative COVID-19 deaths from its effect on cumulative COVID-19 infections. Provided that the cumulative COVID-19 infections follow the same model structure as outlined in equation (2), we can express *I_i,t_* via an exponential growth model:

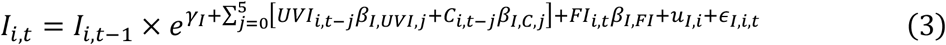

The interpretation of the coefficients and variables is essentially the same as in equation (2) but relates to the cumulative COVID-19 infections rather than deaths. Dividing equation (1) by equation (3) leads to equation (4).

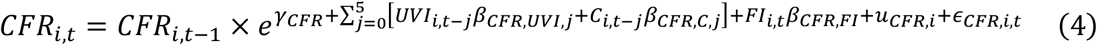

After applying the same transformation on equation (4) to derive equation (2) we get the estimable equation (5).

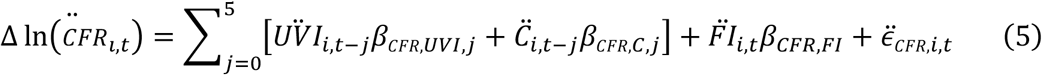

Every coefficient of equation (5) shows the effect of UVI or the effect of a control variable on CFR. For example, *β_CFR,UVI,j_* stands for the effect of UVI lagged by *j* weeks on CFR. Moreover, *β_CFR,UVI,j_* reflects the difference of the effect of UVI on the cumulative COVID-19 deaths and infections, because *β_CFR,UVI,j_ =β_UVI,j_ − β_I,UVI,j_*. This relationship also holds for *β_CFR,C,j_* and *β_CFR,FI_*. We expect to find smaller effect sizes for UVI on CFR. On the one hand, we expect UVI to decrease the cumulative COVID-19 deaths. On the other hand, we expect UVI to decrease the cumulative COVID-19 infections. The reason is not that fewer people get infected but rather that the infections are less severe which makes it less likely that people get themselves tested. One concern when using the observed case fatality rate *CFR^obs^* that is obtained during an epidemic, is that it likely understates the true case fatality rate *CFR^true^*. Note, however, that the model is robust to a miss-reported value of *CFR^true^* as long as the error is multiplicative and time-constant. Suppose the observed case fatality rate

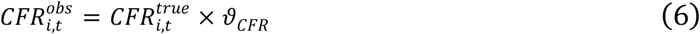

where *ϑ_CFR_* represents the multiplicative and time-constant error. This relationship could be the result of miss-reported cumulative COVID-19 deaths and infections, if there are time-constant errors for the cumulative COVID-19 deaths and infections, because

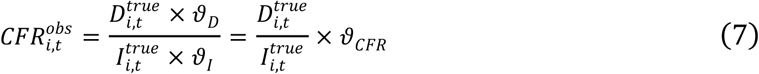

where *ϑ_D_* and *ϑ_I_* represent the multiplicative and time-constant error for the cumulative COVID-19 deaths and infections, respectively. If, for example, the true cumulative COVID-19 deaths at day t are always 10% higher than the observed one, then this 10% difference represents a multiplicative and time-constant error.

If the multiplicative errors of cumulative COVID-19 deaths and infections are not time-constant but grow linearly over time, then the multiplicative error of CFR will still be time constant:

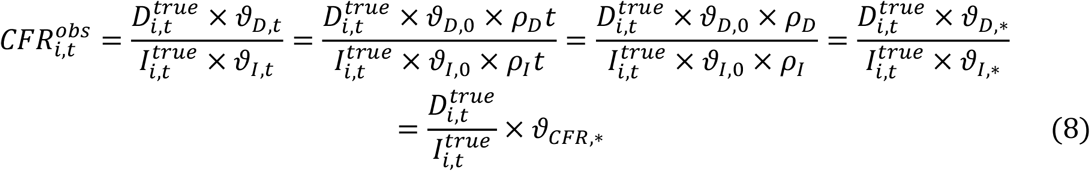

where *ϑ_D,t_* and *ϑ_I,t_* represent the multiplicative error for the cumulative COVID-19 deaths and infections, respectively, which could grow linearly over time. For example, if the true cumulative Covid-19 deaths or infections are first underreported and later over-reported, then *ρ_D_* or *ρ_I_* will be positive and *ϑ_D,t_* as well as *ϑ_I,t_* will first be negative and later (i.e., with larger values for *t*) become positive.

Despite those two forms of measurement-error outlined in equations (7) and (8), our statistical analysis is able to identify the effect of UVI on CFR.

The four aforementioned changes in growth rates of cumulative COVID-19 deaths with respect to time threaten a causal identification of the effect of UVI. Therefore, in addition to controlling for time-constant country-specific changes of growth rates as well as linear changes of growth rates that are similar across countries, we increase the flexibility of our main model specification. The more flexible model isolates the effect of UVI from time trends by allowing time to affect the growth rates of all countries linearly or non-linearly in the same or different way. Essentially, in addition to *FI_i,t_* we also include (*FI_i,t_*)*^2^* and 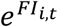 into the model, and we interact each of the variables *FI_i,t_*, (*FI_i,t_*)*^2^* and 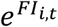 with 64 dummy variables, one for each country, to allow for country-specific linear and non-linear time effects.

### 13.6 Derivation of Equation (2)

Start from equation (1).

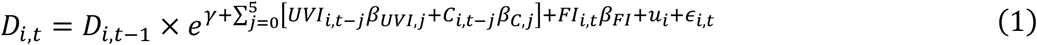

Taking the natural logarithm leads to equation (1.1).

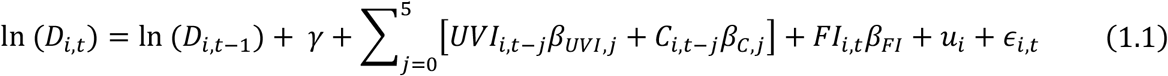

Deducting *ln* (*D_i,t−1_*) leads to equation (1.2)

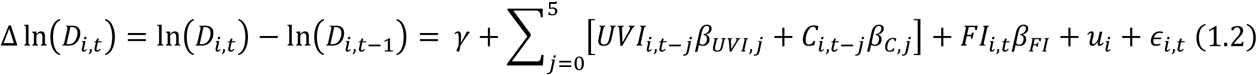

The average version of the left-hand side and right-hand side of equation (1.2) across time is given by equation (1.3) and (1.4), respectively.

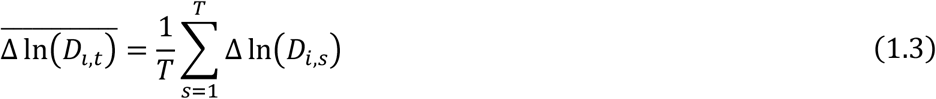

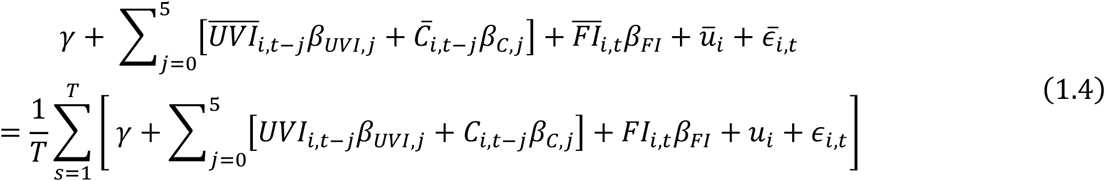

Deducting equation (1.3) and (1.4) from equation (1.2) leads to equation (2).

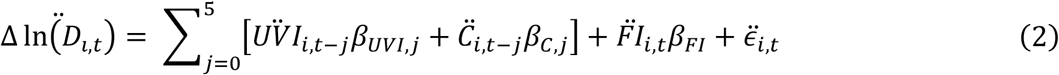

### 13.7 Identification of Daily Growth Rate *γ*

Instead of demeaning equation (1) we can assume that *u_i_* is uncorrelated to all explanatory variables such that a random effects model can be estimated. Under these more restrictive assumptions, *γ* is identified. The results are provided in Table S4. The daily growth rate is estimated to increase COVID-19 deaths by 61.98% [*e*^0.4823^−1] each day. A robust Hausman test to assess the plausibility of the additional assumptions required to identify *γ* is not rejected. Therefore, a random effects model can be used. Nevertheless, the estimate needs to be interpreted with caution because the magnitude lacks theoretical plausibility.

### 13.8 Extended Regression Results (Fixed Effects Analysis)

Table S5, S6, and S7 outline the estimation results of our main model up to 8 weeks lagged for different sets of control variables. The estimates do not change substantially after we include three or more lags of UVI or the control variables. We find that the model with three lags is favorable and we use this model in our main analysis.

### 13.9 Supplementary Tables

**Table S1:** Short- and Long-Run Effects of One-Unit Increase in Ultraviolet Index (UVI) at Time

| Weeks Passed by | Change of $D_{t+s}$ |
| --- | --- |
| 0 | $\beta_{UV,0} \times 100\%$ |
| 1 | $(\beta_{UV,0} + \beta_{UV,1}) \times 100\%$ |
| 2 | $(\beta_{UV,0} + \beta_{UV,1} + \beta_{UV,2}) \times 100\%$ |
| 3 | $(\beta_{UV,0} + \beta_{UV,1} + \beta_{UV,2} + \beta_{UV,3}) \times 100\%$ |
| 4 | $(\beta_{UV,0} + \beta_{UV,1} + \beta_{UV,2} + \beta_{UV,3} + \beta_{UV,4}) \times 100\%$ |
| 5 or more | $(\beta_{UV,0} + \beta_{UV,1} + \beta_{UV,2} + \beta_{UV,3} + \beta_{UV,4} + \beta_{UV,5}) \times 100\%$ |

**Table S2:** Robustness Check for Linear and Quadratic Time Trends

|  | Model 3 | Model 4 | Model 5 |
| --- | --- | --- | --- |
| Dependent Variables | COVID-19 Deaths | COVID-19 Deaths | COVID-19 Deaths |
| L0.UVI | -0.002 (-1.53) | -0.000 (-0.24) | 0.000 (0.05) |
| L1.UVI | 0.000 (0.08) | 0.002 (0.92) | 0.001 (0.43) |
| L2.UVI | -0.001 (-0.97) | -0.000 (-0.28) | -0.001 (-0.45) |
| L3.UVI | -0.002 (-1.36) | -0.002 (-1.10) | -0.002 (-1.33) |
| L4.UVI | -0.004* (-2.08) | -0.003+ (-1.94) | -0.004* (-2.02) |
| L5.UVI | -0.002 (-1.29) | -0.002 (-0.99) | -0.002 (-1.13) |
| Long-Run Coefficient | -0.012** (F: 8.76) | -0.006 (F: 2.45) | -0.007* (F: 4.04) |
| **Control Variables** |  |  |  |
| Time Trend | Linear and Square | Linear<br>(country-specific) | Linear and Square<br>(country-specific) |
| Country Fixed Effects | Yes | Yes | Yes |
| Precipitation index | Yes | Yes | Yes |
| Cloud index | Yes | Yes | Yes |
| Ozone level | Yes | Yes | Yes |
| Visibility Level | Yes | Yes | Yes |
| Humidity level | Yes | Yes | Yes |
| Temperature (min and max) | Yes | Yes | Yes |
| Number of Estimates | 50 (+152 FE) | 48 (+152 FE + 152 TSCE) | 48 (+152 FE + 304 TSCE) |
| Number of Observations | 6,524 | 6,524 | 6,524 |
| Number of Countries | 152 | 152 | 152 |
| R-squared Within | 14.00% | 19.01% | 24.04% |

Note: +: $p < 0.10$ , \*: $p < 0.05$ , \*\*: $p < 0.01$ . t-statistics based on robust standard errors in parentheses. F-statistic for long-run coefficient in parentheses. L0.UVI stands for the effect of UVI at time $t$ on the cumulative number of COVID-19 deaths at the same time, whereas L1.UVI, L2.UVI, L3.UVI, L4.UVI and L5.UVI stand for the effect of UVI lagged by one, two, or three weeks respectively. FE stands for country fixed-effects, TSCE stands for time country-specific effects.

**Table S3:** Robustness Check for Exponential, Linear, Quadratic Time Trends

|  | Model 6 | Model 7 | Model 8 |
| --- | --- | --- | --- |
| Dependent Variables | COVID-19 Deaths | COVID-19 Deaths | COVID-19 Deaths |
| L0.UVI | -0.003 (-1.60) | -0.000 (-0.12) | -0.000 (-0.25) |
| L1.UVI | 0.000 (0.01) | 0.002 (0.91) | 0.001 (0.44) |
| L2.UVI | -0.001 (-0.95) | -0.001 (-0.42) | -0.001 (-0.53) |
| L3.UVI | -0.002 (-1.28) | -0.002 (-1.06) | -0.002 (-1.39) |
| L4.UVI | -0.004* (-2.06) | -0.003+ (-1.82) | -0.004+ (-1.91) |
| L5.UVI | -0.002 (-1.25) | -0.002 (-1.00) | -0.002 (-1.15) |
| Long-Run Coefficient | -0.012** (F: 8.53) | -0.006 (F2.27) | -0.008* (F: 4.69) |
| **Control Variables** |  |  |  |
| Time Trend | Linear and exponential | Linear and exponential (country-specific) | Linear, Square and exponential (country-specific) |
| Country Fixed Effects | Yes | Yes | Yes |
| Precipitation index | Yes | Yes | Yes |
| Cloud index | Yes | Yes | Yes |
| Ozone level | Yes | Yes | Yes |
| Visibility Level | Yes | Yes | Yes |
| Humidity level | Yes | Yes | Yes |
| Temperature (min and max) | Yes | Yes | Yes |
| Number of Estimates | 50 (+152 FE) | 48 (+152 FE + 304 TSCE) | 48 (+152 FE + 456 TSCE) |
| Number of Observations | 6,524 | 6,524 | 6,524 |
| Number of Countries | 152 | 152 | 152 |
| R-squared Within | 13.78% | 19.64% | 25.00% |
| Note: +: $p < 0.10$, *: $p < 0.05$, **: $p < 0.01$. t-statistics based on robust standard errors in parentheses. F-statistic for long-run coefficient in parentheses. L0.UVI stands for the effect of UVI at time t on the cumulative number of COVID-19 deaths at the same time, whereas L1.UVI, L2.UVI, L3.UVI, L4.UVI and L5.UVI stand for the effect of UVI lagged by one, two, or three weeks respectively. FE stands for country fixed-effects, TCSE stands for time country-specific effects. | | | |

**Table S4:** Identification of Daily Growth Rate *γ*

|  | Model 9 |
| --- | --- |
| $\gamma$ | 0.4823 |
| L0.UVI | -0.002 (-0.93) |
| L1.UVI | 0.000 (0.17) |
| L2.UVI | -0.001 (-0.99) |
| L3.UVI | -0.002 (-1.57) |
| L4.UVI | -0.004* (-2.45) |
| L5.UVI | -0.004* (-2.14) |
| Long-Run Coefficient | -0.013*** (F: 13.63) |
| Control Variables |  |
| Time Trend | Linear |
| Precipitation index | Yes |
| Cloud index | Yes |
| Ozone level | Yes |
| Visibility Level | Yes |
| Humidity level | Yes |
| Temperature (min and max) | Yes |
| Number of Estimates | 50 |
| Number of Observations | 6,524 |
| Number of Countries | 152 |
| R-squared within | 13.03% |
| p-value of Robust Hausman-test | 0.6661 |

Note: +: $p < 0.10$ , \*: $p < 0.05$ , \*\*: $p < 0.01$ . t-statistics based on robust standard errors in parentheses. F-statistic for long-run coefficient in parentheses. Robust Hausman-test based on 2,000 clustered bootstrap replications. L0.UVI stands for the effect of UVI at time $t$ on the cumulative number of COVID-19 deaths at the same time, whereas L1.UVI, L2.UVI, L3.UVI, L4.UVI and L5.UVI stand for the effect of UVI lagged by one, two, three, four or five weeks respectively. FE stands for country fixed-effects.

**Table S5:** Estimation of Models with Different Lags without Control Variables

|  | Model<br>A1.1 | Model<br>A1.2 | Model<br>A1.3 | Model<br>A1.4 | Model<br>A1.5 | Model<br>A1.6 | Model<br>A1.7 | Model<br>A1.8 | Model<br>A1.9 |
| --- | --- | --- | --- | --- | --- | --- | --- | --- | --- |
| Time Since<br>First Infection | -0.003***<br>(-11.53) | -0.003***<br>(-11.48) | -0.003***<br>(-11.33) | -0.003***<br>(-11.05) | -0.003***<br>(-11.95) | -0.004***<br>(-13.76) | -0.004***<br>(-14.80) | -0.004***<br>(-16.61) | -0.004***<br>(-17.04) |
| L0.UVI | -0.001 (-<br>1.21) | -0.001 (-<br>1.22) | -0.001 (-<br>1.19) | -0.001 (-<br>1.10) | -0.001 (-<br>1.15) | -0.001 (-<br>1.04) | -0.001 (-<br>0.88) | -0.001 (-<br>0.88) | -0.001 (-<br>0.97) |
| L1.UVI |  | 0.000<br>(0.04) | 0.000<br>(0.09) | 0.000<br>(0.14) | 0.000<br>(0.11) | 0.000<br>(0.34) | 0.001<br>(0.50) | 0.001<br>(0.47) | 0.001<br>(0.95) |
| L2.UVI |  |  | -0.002 (-<br>1.51) | -0.001 (-<br>1.40) | -0.001 (-<br>1.31) | -0.001 (-<br>1.18) | -0.001 (-<br>0.97) | -0.001 (-<br>0.74) | -0.001 (-<br>0.54) |
| L3.UVI |  |  |  | -0.003**<br>(-2.98) | -0.003**<br>(-3.01) | -0.003**<br>(-2.91) | -0.003**<br>(-2.82) | -0.003**<br>(-2.83) | -0.003**<br>(-2.62) |
| L4.UVI |  |  |  |  | -0.001 (-<br>0.84) | -0.001 (-<br>0.98) | -0.001 (-<br>1.00) | -0.001 (-<br>1.00) | -0.001 (-<br>1.13) |
| L5.UVI |  |  |  |  |  | -0.000 (-<br>0.28) | -0.000 (-<br>0.43) | -0.001 (-<br>0.51) | -0.001 (-<br>0.57) |
| L6.UVI |  |  |  |  |  |  | -0.000 (-<br>0.17) | -0.000 (-<br>0.32) | -0.000 (-<br>0.09) |
| L7.UVI |  |  |  |  |  |  |  | -0.002* (-<br>2.16) | -0.002* (-<br>2.11) |
| L8.UVI |  |  |  |  |  |  |  |  | -0.001 (-<br>0.93) |
| Constant | 0.261***<br>(15.24) | 0.261***<br>(14.02) | 0.268***<br>(13.49) | 0.282***<br>(13.93) | 0.291***<br>(14.13) | 0.297***<br>(14.94) | 0.299***<br>(15.10) | 0.313***<br>(15.65) | 0.312***<br>(15.39) |
| Number of<br>Observations | 6,588 | 6,588 | 6,588 | 6,586 | 6,568 | 6,524 | 6,468 | 6,381 | 6,223 |
| Number of<br>States | 152 | 152 | 152 | 152 | 152 | 152 | 152 | 152 | 152 |
| Adjusted R-squared | 0.116 | 0.116 | 0.116 | 0.117 | 0.121 | 0.125 | 0.128 | 0.130 | 0.130 |

Note: +: $p < 0.10$ , \*: $p < 0.05$ , \*\*: $p < 0.01$ . t-statistics based on robust standard errors in parentheses. L0.UVI stands for the effect of UVI at time $t$ on the cumulative number of COVID-19 deaths at the same time, whereas L1.UVI, L2.UVI and L3.UVI (etc.) stand for the effect of UVI lagged by one, two, or three (etc.) weeks respectively. The same is true for the other variables.

**Table S6:** Estimation of Models with Different Lags with Control Variables Affecting Ultraviolet Index

|  | Model<br>A2.1 | Model<br>A2.2 | Model<br>A2.3 | Model<br>A2.4 | Model<br>A2.5 | Model<br>A2.6 | Model<br>A2.7 | Model<br>A2.8 | Model<br>A2.9 |
| --- | --- | --- | --- | --- | --- | --- | --- | --- | --- |
| Time Since First<br>Infection | -<br>0.003***<br>(-11.31) | -<br>0.003***<br>(-11.20) | -<br>0.003***<br>(-11.13) | -<br>0.003***<br>(-11.16) | -<br>0.003***<br>(-12.12) | -<br>0.003***<br>(-13.38) | -<br>0.004***<br>(-14.34) | -<br>0.004***<br>(-15.10) | -<br>0.004***<br>(-14.57) |
| L0.UVI | -0.002 (-<br>1.42) | -0.002 (-<br>1.34) | -0.002 (-<br>1.25) | -0.002 (-<br>1.24) | -0.002 (-<br>1.34) | -0.002 (-<br>1.24) | -0.002 (-<br>1.18) | -0.002 (-<br>1.06) | -0.002 (-<br>0.96) |
| L1.UVI |  | 0.000<br>(0.27) | 0.001<br>(0.32) | 0.001<br>(0.34) | 0.001<br>(0.32) | 0.001<br>(0.43) | 0.001<br>(0.39) | 0.001<br>(0.51) | 0.001<br>(0.72) |
| L2.UVI |  |  | -0.000 (-<br>0.29) | -0.000 (-<br>0.22) | -0.000 (-<br>0.21) | -0.000 (-<br>0.07) | -0.000 (-<br>0.18) | 0.000<br>(0.28) | 0.000<br>(0.23) |
| L3.UVI |  |  |  | -0.001 (-<br>0.39) | -0.000 (-<br>0.18) | -0.001 (-<br>0.44) | -0.001 (-<br>0.39) | -0.001 (-<br>0.44) | -0.000 (-<br>0.17) |
| L4.UVI |  |  |  |  | -0.002 (-<br>1.19) | -0.002 (-<br>1.34) | -0.002 (-<br>1.44) | -0.002 (-<br>1.48) | -0.003+<br>(-1.81) |
| L5.UVI |  |  |  |  |  | -0.001 (-<br>0.68) | -0.001 (-<br>0.70) | -0.002 (-<br>1.08) | -0.002 (-<br>1.02) |
| L6.UVI |  |  |  |  |  |  | 0.001<br>(0.72) | 0.001<br>(0.64) | 0.001<br>(0.57) |
| L7.UVI |  |  |  |  |  |  |  | -0.002 (-<br>1.46) | -0.002 (-<br>1.38) |
| L8.UVI |  |  |  |  |  |  |  |  | -0.000 (-<br>0.12) |
| L0.CLOUD | -0.014 (-<br>1.16) | -0.014 (-<br>1.12) | -0.013 (-<br>1.01) | -0.014 (-<br>1.12) | -0.016 (-<br>1.25) | -0.015 (-<br>1.17) | -0.014 (-<br>1.08) | -0.012 (-<br>0.96) | -0.012 (-<br>0.92) |
| L1.CLOUD |  | -0.009 (-<br>0.75) | -0.007 (-<br>0.62) | -0.007 (-<br>0.59) | -0.006 (-<br>0.56) | -0.008 (-<br>0.72) | -0.009 (-<br>0.83) | -0.006 (-<br>0.51) | -0.007 (-<br>0.60) |
| L2.CLOUD |  |  | 0.002<br>(0.22) | 0.003<br>(0.31) | 0.004<br>(0.34) | 0.004<br>(0.32) | 0.001<br>(0.08) | 0.006<br>(0.52) | 0.005<br>(0.45) |
| L3.CLOUD |  |  |  | 0.014<br>(1.35) | 0.017<br>(1.63) | 0.012<br>(1.18) | 0.012<br>(1.24) | 0.011<br>(1.13) | 0.016+<br>(1.75) |
| L4.CLOUD |  |  |  |  | -0.004 (-<br>0.33) | -0.005 (-<br>0.44) | -0.004 (-<br>0.30) | -0.004 (-<br>0.36) | -0.007 (-<br>0.65) |
| L5.CLOUD |  |  |  |  |  | -0.006 (-<br>0.55) | -0.003 (-<br>0.27) | -0.010 (-<br>0.89) | -0.007 (-<br>0.65) |
| L6.CLOUD |  |  |  |  |  |  | 0.008<br>(0.76) | 0.009<br>(0.93) | 0.008<br>(0.86) |
| L7.CLOUD |  |  |  |  |  |  |  | -0.002 (-<br>0.17) | 0.002<br>(0.16) |
| L8.CLOUD |  |  |  |  |  |  |  |  | 0.013<br>(1.46) |
| L0.PRECIP | -0.016+<br>(-1.73) | -0.015 (-<br>1.63) | -0.015 (-<br>1.65) | -0.013 (-<br>1.46) | -0.011 (-<br>1.23) | -0.011 (-<br>1.16) | -0.010 (-<br>1.12) | -0.008 (-<br>0.87) | -0.007 (-<br>0.73) |
| L1.PRECIP |  | 0.019+<br>(1.83) | 0.019+<br>(1.79) | 0.019+<br>(1.80) | 0.018+<br>(1.77) | 0.023*<br>(2.20) | 0.020*<br>(2.10) | 0.017+<br>(1.78) | 0.016+<br>(1.80) |
| L2.PRECIP |  |  | 0.005<br>(0.45) | 0.003<br>(0.28) | 0.003<br>(0.24) | 0.004<br>(0.40) | 0.006<br>(0.51) | 0.006<br>(0.54) | 0.006<br>(0.53) |
| L3.PRECIP |  |  |  | 0.018<br>(1.54) | 0.018<br>(1.54) | 0.019+<br>(1.67) | 0.018<br>(1.56) | 0.018<br>(1.64) | 0.016<br>(1.35) |
| L4.PRECIP |  |  |  |  | -0.009 (-<br>1.02) | -0.008 (-<br>1.01) | -0.010 (-<br>1.24) | -0.009 (-<br>1.08) | -0.009 (-<br>1.05) |
| L5.PRECIP |  |  |  |  |  | -0.005 (-<br>0.57) | -0.006 (-<br>0.72) | -0.005 (-<br>0.57) | -0.013 (-<br>1.54) |
| L6.PRECIP |  |  |  |  |  |  | 0.001<br>(0.16) | -0.002 (-<br>0.26) | -0.004 (-<br>0.45) |
| L7.PRECIP |  |  |  |  |  |  |  | 0.008<br>(0.87) | 0.004<br>(0.46) |
| L8.PRECIP |  |  |  |  |  |  |  |  | 0.007<br>(0.69) |
| L0.OZONE | 0.000+<br>(1.91) | 0.000+<br>(1.77) | 0.000+<br>(1.77) | 0.000<br>(1.46) | 0.000<br>(1.17) | 0.000<br>(1.13) | 0.000<br>(1.30) | 0.000<br>(1.24) | 0.000<br>(1.48) |
| L1.OZONE |  | 0.000<br>(0.76) | 0.000<br>(0.56) | 0.000<br>(0.41) | 0.000<br>(0.58) | 0.000<br>(0.69) | 0.000<br>(1.36) | 0.000<br>(1.38) | 0.000<br>(1.62) |
| L2.OZONE |  |  | 0.000<br>(0.49) | 0.000<br>(0.33) | -0.000 (-<br>0.01) | -0.000 (-<br>0.10) | -0.000 (-<br>0.36) | -0.000 (-<br>0.57) | -0.000 (-<br>1.17) |
| L3.OZONE |  |  |  | -0.000 (-<br>0.13) | 0.000<br>(0.08) | 0.000<br>(0.15) | 0.000<br>(0.56) | 0.000<br>(0.58) | 0.000<br>(0.18) |
| L4.OZONE |  |  |  |  | -0.000 (-<br>0.67) | -0.000 (-<br>0.73) | -0.000 (-<br>0.84) | -0.000 (-<br>1.07) | -0.000 (-<br>1.23) |
| L5.OZONE |  |  |  |  |  | -0.000 (-<br>0.34) | -0.000 (-<br>0.62) | -0.000 (-<br>0.27) | 0.000<br>(0.61) |
| L6.OZONE |  |  |  |  |  |  | 0.000*<br>(2.18) | 0.000+<br>(1.97) | 0.000<br>(1.36) |
| L7.OZONE |  |  |  |  |  |  |  | -0.000 (-<br>0.84) | -0.000 (-<br>0.81) |
| L8.OZONE |  |  |  |  |  |  |  |  | -0.000* (-<br>2.10) |
| L0.VISIBILITY | -0.003* (-<br>2.29) | -0.003* (-<br>2.26) | -0.003* (-<br>2.30) | -0.003* (-<br>2.26) | -0.003* (-<br>2.24) | -0.003* (-<br>2.03) | -0.002 (-<br>1.61) | -0.002 (-<br>1.57) | -0.003+<br>(-1.84) |
| L1.VISIBILITY |  | 0.001<br>(0.54) | 0.001<br>(0.57) | 0.000<br>(0.44) | 0.001<br>(0.58) | 0.001<br>(1.28) | 0.001<br>(1.11) | 0.001<br>(1.11) | 0.001<br>(1.22) |
| L2.VISIBILITY |  |  | -0.001 (-<br>0.77) | -0.001 (-<br>0.97) | -0.001 (-<br>0.85) | -0.001 (-<br>0.70) | -0.001 (-<br>0.54) | -0.000 (-<br>0.39) | -0.000 (-<br>0.20) |
| L3.VISIBILITY |  |  |  | 0.000<br>(0.18) | 0.000<br>(0.28) | 0.001<br>(0.46) | 0.001<br>(0.54) | 0.000<br>(0.42) | 0.001<br>(0.92) |
| L4.VISIBILITY |  |  |  |  | -0.000 (-<br>0.01) | -0.000 (-<br>0.09) | -0.000 (-<br>0.02) | 0.000<br>(0.11) | 0.000<br>(0.23) |
| L5.VISIBILITY |  |  |  |  |  | -0.001 (-<br>0.62) | -0.000 (-<br>0.47) | -0.001 (-<br>0.59) | -0.001 (-<br>1.08) |
| L6.VISIBILITY |  |  |  |  |  |  | 0.000<br>(0.09) | -0.000 (-<br>0.23) | 0.000<br>(0.02) |
| L7.VISIBILITY |  |  |  |  |  |  |  | -0.001 (-<br>0.55) | -0.001 (-<br>0.54) |
| L8.VISIBILITY |  |  |  |  |  |  |  |  | 0.001<br>(1.06) |
| Constant | 0.242***<br>(4.22) | 0.208*<br>(2.38) | 0.209+<br>(1.81) | 0.227<br>(1.58) | 0.268<br>(1.65) | 0.290+<br>(1.87) | 0.193<br>(1.22) | 0.255+<br>(1.67) | 0.293+<br>(1.71) |
| Number of Observations | 6,588 | 6,588 | 6,588 | 6,586 | 6,568 | 6,524 | 6,468 | 6,381 | 6,223 |
| Number of States | 152 | 152 | 152 | 152 | 152 | 152 | 152 | 152 | 152 |
| Adjusted R-squared | 0.118 | 0.119 | 0.120 | 0.122 | 0.126 | 0.131 | 0.134 | 0.136 | 0.137 |

Note: +: $p < 0.10$ , \*: $p < 0.05$ , \*\*: $p < 0.01$ . t-statistics based on robust standard errors in parentheses. L0.UVI stands for the effect of UVI at time t on the cumulative number of COVID-19 deaths at the same time, whereas L1.UVI, L2.UVI and L3.UVI (etc.) stand for the effect of UVI lagged by one, two, or three (etc.) weeks respectively. The same is true for the other variables.

**Table S7:**
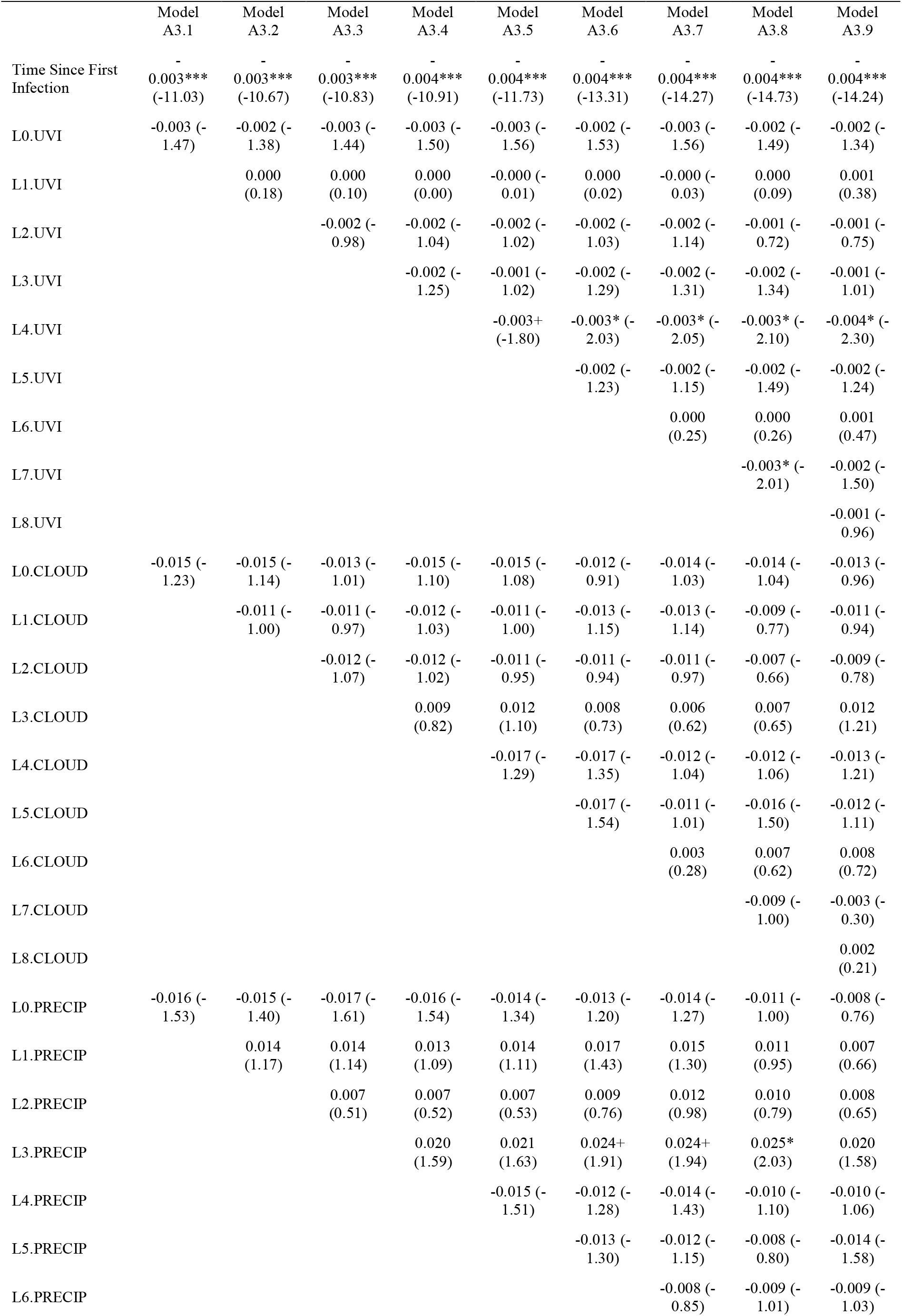
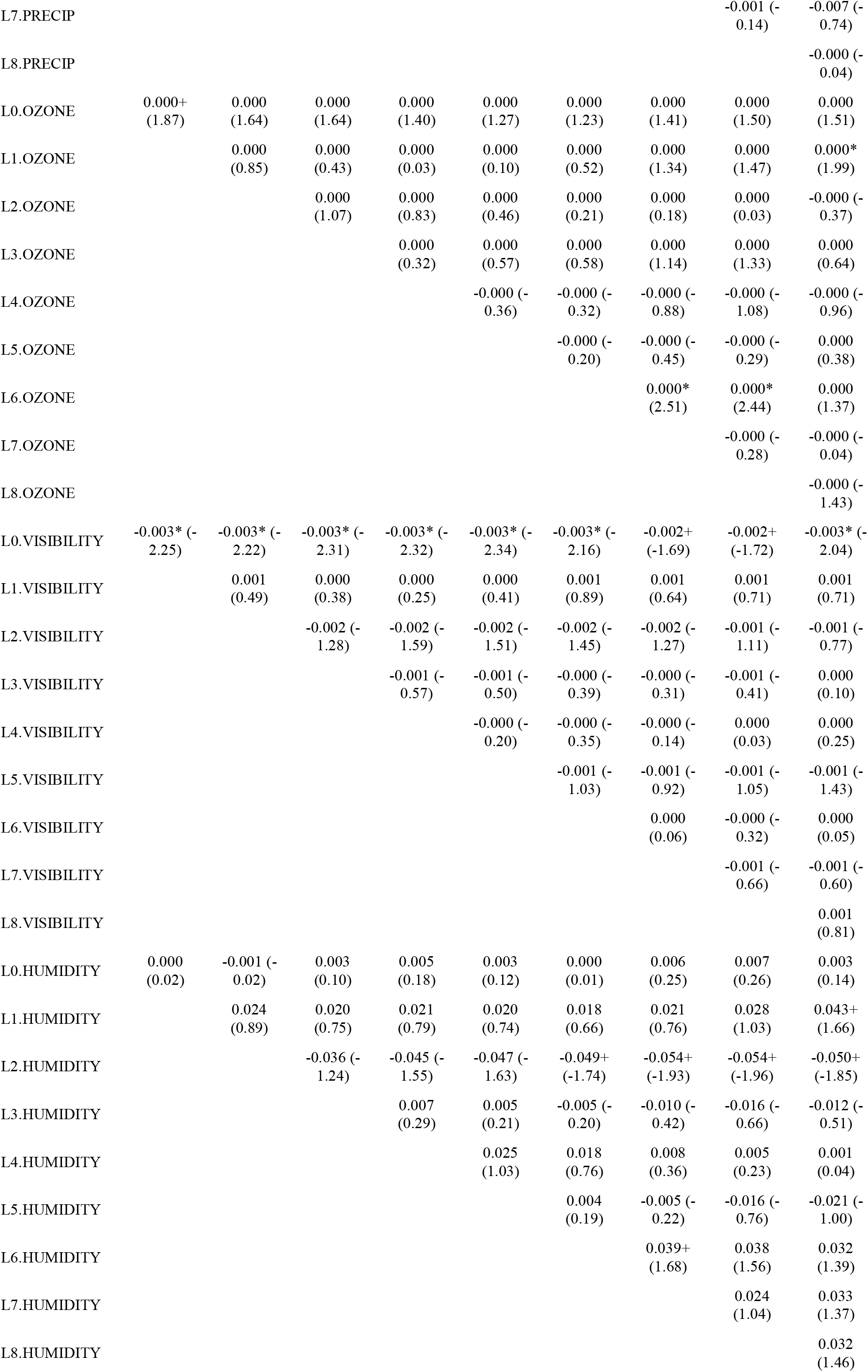
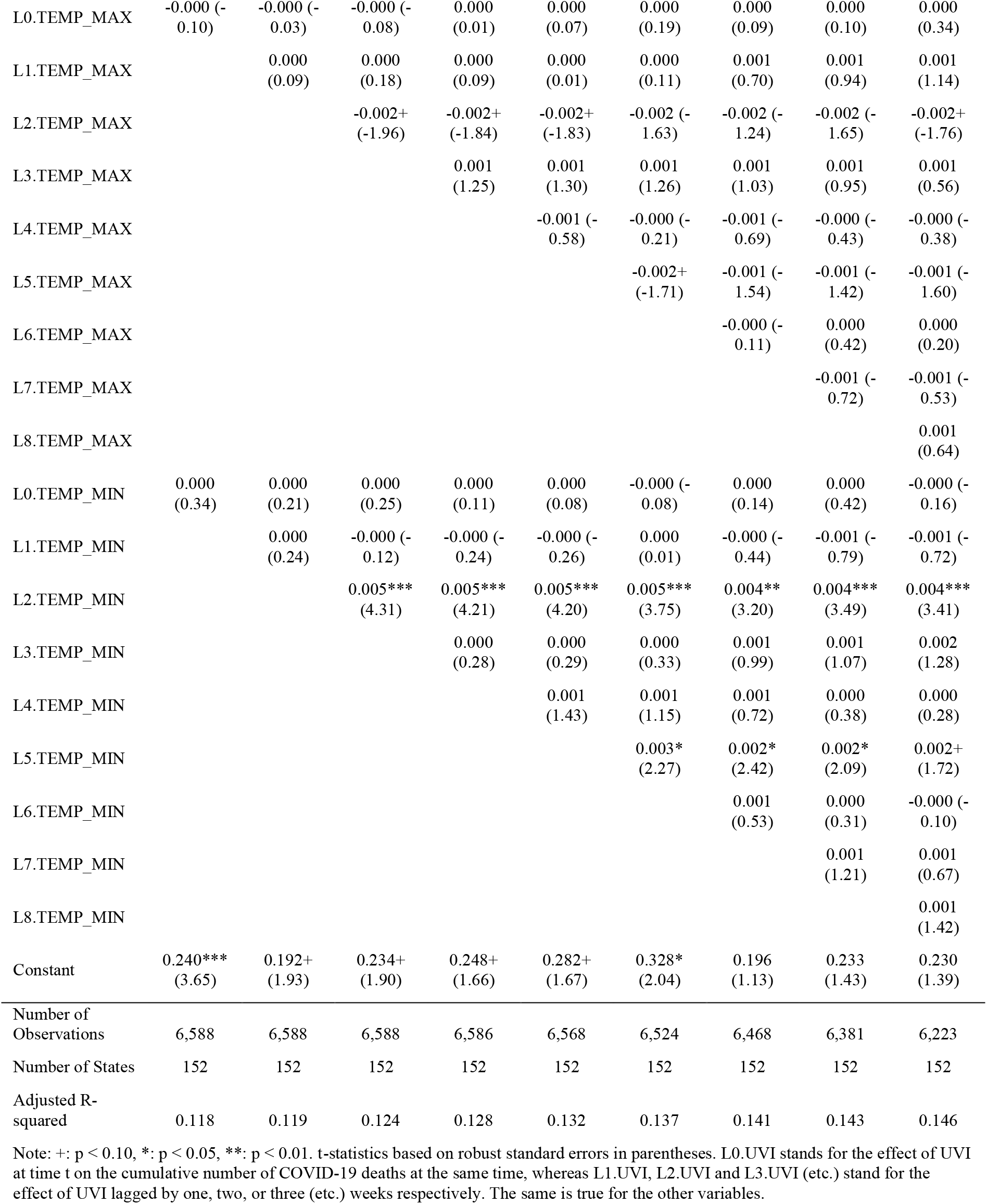
Estimation of Models with Different Lags with All Control Variables

|  | Model<br>A3.1 | Model<br>A3.2 | Model<br>A3.3 | Model<br>A3.4 | Model<br>A3.5 | Model<br>A3.6 | Model<br>A3.7 | Model<br>A3.8 | Model<br>A3.9 |
| --- | --- | --- | --- | --- | --- | --- | --- | --- | --- |
| Time Since First<br>Infection | -<br>0.003***<br>(-11.03) | -<br>0.003***<br>(-10.67) | -<br>0.003***<br>(-10.83) | -<br>0.004***<br>(-10.91) | -<br>0.004***<br>(-11.73) | -<br>0.004***<br>(-13.31) | -<br>0.004***<br>(-14.27) | -<br>0.004***<br>(-14.73) | -<br>0.004***<br>(-14.24) |
| L0.UVI | -0.003 (-<br>1.47) | -0.002 (-<br>1.38) | -0.003 (-<br>1.44) | -0.003 (-<br>1.50) | -0.003 (-<br>1.56) | -0.002 (-<br>1.53) | -0.003 (-<br>1.56) | -0.002 (-<br>1.49) | -0.002 (-<br>1.34) |
| L1.UVI |  | 0.000<br>(0.18) | 0.000<br>(0.10) | 0.000<br>(0.00) | -0.000 (-<br>0.01) | 0.000<br>(0.02) | -0.000 (-<br>0.03) | 0.000<br>(0.09) | 0.001<br>(0.38) |
| L2.UVI |  |  | -0.002 (-<br>0.98) | -0.002 (-<br>1.04) | -0.002 (-<br>1.02) | -0.002 (-<br>1.03) | -0.002 (-<br>1.14) | -0.001 (-<br>0.72) | -0.001 (-<br>0.75) |
| L3.UVI |  |  |  | -0.002 (-<br>1.25) | -0.001 (-<br>1.02) | -0.002 (-<br>1.29) | -0.002 (-<br>1.31) | -0.002 (-<br>1.34) | -0.001 (-<br>1.01) |
| L4.UVI |  |  |  |  | -0.003+<br>(-1.80) | -0.003* (-<br>2.03) | -0.003* (-<br>2.05) | -0.003* (-<br>2.10) | -0.004* (-<br>2.30) |
| L5.UVI |  |  |  |  |  | -0.002 (-<br>1.23) | -0.002 (-<br>1.15) | -0.002 (-<br>1.49) | -0.002 (-<br>1.24) |
| L6.UVI |  |  |  |  |  |  | 0.000<br>(0.25) | 0.000<br>(0.26) | 0.001<br>(0.47) |
| L7.UVI |  |  |  |  |  |  |  | -0.003* (-<br>2.01) | -0.002 (-<br>1.50) |
| L8.UVI |  |  |  |  |  |  |  |  | -0.001 (-<br>0.96) |
| L0.CLOUD | -0.015 (-<br>1.23) | -0.015 (-<br>1.14) | -0.013 (-<br>1.01) | -0.015 (-<br>1.10) | -0.015 (-<br>1.08) | -0.012 (-<br>0.91) | -0.014 (-<br>1.03) | -0.014 (-<br>1.04) | -0.013 (-<br>0.96) |
| L1.CLOUD |  | -0.011 (-<br>1.00) | -0.011 (-<br>0.97) | -0.012 (-<br>1.03) | -0.011 (-<br>1.00) | -0.013 (-<br>1.15) | -0.013 (-<br>1.14) | -0.009 (-<br>0.77) | -0.011 (-<br>0.94) |
| L2.CLOUD |  |  | -0.012 (-<br>1.07) | -0.012 (-<br>1.02) | -0.011 (-<br>0.95) | -0.011 (-<br>0.94) | -0.011 (-<br>0.97) | -0.007 (-<br>0.66) | -0.009 (-<br>0.78) |
| L3.CLOUD |  |  |  | 0.009<br>(0.82) | 0.012<br>(1.10) | 0.008<br>(0.73) | 0.006<br>(0.62) | 0.007<br>(0.65) | 0.012<br>(1.21) |
| L4.CLOUD |  |  |  |  | -0.017 (-<br>1.29) | -0.017 (-<br>1.35) | -0.012 (-<br>1.04) | -0.012 (-<br>1.06) | -0.013 (-<br>1.21) |
| L5.CLOUD |  |  |  |  |  | -0.017 (-<br>1.54) | -0.011 (-<br>1.01) | -0.016 (-<br>1.50) | -0.012 (-<br>1.11) |
| L6.CLOUD |  |  |  |  |  |  | 0.003<br>(0.28) | 0.007<br>(0.62) | 0.008<br>(0.72) |
| L7.CLOUD |  |  |  |  |  |  |  | -0.009 (-<br>1.00) | -0.003 (-<br>0.30) |
| L8.CLOUD |  |  |  |  |  |  |  |  | 0.002<br>(0.21) |
| L0.PRECIP | -0.016 (-<br>1.53) | -0.015 (-<br>1.40) | -0.017 (-<br>1.61) | -0.016 (-<br>1.54) | -0.014 (-<br>1.34) | -0.013 (-<br>1.20) | -0.014 (-<br>1.27) | -0.011 (-<br>1.00) | -0.008 (-<br>0.76) |
| L1.PRECIP |  | 0.014<br>(1.17) | 0.014<br>(1.14) | 0.013<br>(1.09) | 0.014<br>(1.11) | 0.017<br>(1.43) | 0.015<br>(1.30) | 0.011<br>(0.95) | 0.007<br>(0.66) |
| L2.PRECIP |  |  | 0.007<br>(0.51) | 0.007<br>(0.52) | 0.007<br>(0.53) | 0.009<br>(0.76) | 0.012<br>(0.98) | 0.010<br>(0.79) | 0.008<br>(0.65) |
| L3.PRECIP |  |  |  | 0.020<br>(1.59) | 0.021<br>(1.63) | 0.024+<br>(1.91) | 0.024+<br>(1.94) | 0.025*<br>(2.03) | 0.020<br>(1.58) |
| L4.PRECIP |  |  |  |  | -0.015 (-<br>1.51) | -0.012 (-<br>1.28) | -0.014 (-<br>1.43) | -0.010 (-<br>1.10) | -0.010 (-<br>1.06) |
| L5.PRECIP |  |  |  |  |  | -0.013 (-<br>1.30) | -0.012 (-<br>1.15) | -0.008 (-<br>0.80) | -0.014 (-<br>1.58) |
| L6.PRECIP |  |  |  |  |  |  | -0.008 (-<br>0.85) | -0.009 (-<br>1.01) | -0.009 (-<br>1.03) |
| L7.PRECIP |  |  |  |  |  |  |  | -0.001 (-0.14) | -0.007 (-0.74) |
| L8.PRECIP |  |  |  |  |  |  |  |  | -0.000 (-0.04) |
| L0.OZONE | 0.000+ (1.87) | 0.000 (1.64) | 0.000 (1.64) | 0.000 (1.40) | 0.000 (1.27) | 0.000 (1.23) | 0.000 (1.41) | 0.000 (1.50) | 0.000 (1.51) |
| L1.OZONE |  | 0.000 (0.85) | 0.000 (0.43) | 0.000 (0.03) | 0.000 (0.10) | 0.000 (0.52) | 0.000 (1.34) | 0.000 (1.47) | 0.000* (1.99) |
| L2.OZONE |  |  | 0.000 (1.07) | 0.000 (0.83) | 0.000 (0.46) | 0.000 (0.21) | 0.000 (0.18) | 0.000 (0.03) | -0.000 (-0.37) |
| L3.OZONE |  |  |  | 0.000 (0.32) | 0.000 (0.57) | 0.000 (0.58) | 0.000 (1.14) | 0.000 (1.33) | 0.000 (0.64) |
| L4.OZONE |  |  |  |  | -0.000 (-0.36) | -0.000 (-0.32) | -0.000 (-0.88) | -0.000 (-1.08) | -0.000 (-0.96) |
| L5.OZONE |  |  |  |  |  | -0.000 (-0.20) | -0.000 (-0.45) | -0.000 (-0.29) | 0.000 (0.38) |
| L6.OZONE |  |  |  |  |  |  | 0.000* (2.51) | 0.000* (2.44) | 0.000 (1.37) |
| L7.OZONE |  |  |  |  |  |  |  | -0.000 (-0.28) | -0.000 (-0.04) |
| L8.OZONE |  |  |  |  |  |  |  |  | -0.000 (-1.43) |
| L0.VISIBILITY | -0.003* (-2.25) | -0.003* (-2.22) | -0.003* (-2.31) | -0.003* (-2.32) | -0.003* (-2.34) | -0.003* (-2.16) | -0.002+ (-1.69) | -0.002+ (-1.72) | -0.003* (-2.04) |
| L1.VISIBILITY |  | 0.001 (0.49) | 0.000 (0.38) | 0.000 (0.25) | 0.000 (0.41) | 0.001 (0.89) | 0.001 (0.64) | 0.001 (0.71) | 0.001 (0.71) |
| L2.VISIBILITY |  |  | -0.002 (-1.28) | -0.002 (-1.59) | -0.002 (-1.51) | -0.002 (-1.45) | -0.002 (-1.27) | -0.001 (-1.11) | -0.001 (-0.77) |
| L3.VISIBILITY |  |  |  | -0.001 (-0.57) | -0.001 (-0.50) | -0.000 (-0.39) | -0.000 (-0.31) | -0.001 (-0.41) | 0.000 (0.10) |
| L4.VISIBILITY |  |  |  |  | -0.000 (-0.20) | -0.000 (-0.35) | -0.000 (-0.14) | 0.000 (0.03) | 0.000 (0.25) |
| L5.VISIBILITY |  |  |  |  |  | -0.001 (-1.03) | -0.001 (-0.92) | -0.001 (-1.05) | -0.001 (-1.43) |
| L6.VISIBILITY |  |  |  |  |  |  | 0.000 (0.06) | -0.000 (-0.32) | 0.000 (0.05) |
| L7.VISIBILITY |  |  |  |  |  |  |  | -0.001 (-0.66) | -0.001 (-0.60) |
| L8.VISIBILITY |  |  |  |  |  |  |  |  | 0.001 (0.81) |
| L0.HUMIDITY | 0.000 (0.02) | -0.001 (-0.02) | 0.003 (0.10) | 0.005 (0.18) | 0.003 (0.12) | 0.000 (0.01) | 0.006 (0.25) | 0.007 (0.26) | 0.003 (0.14) |
| L1.HUMIDITY |  | 0.024 (0.89) | 0.020 (0.75) | 0.021 (0.79) | 0.020 (0.74) | 0.018 (0.66) | 0.021 (0.76) | 0.028 (1.03) | 0.043+ (1.66) |
| L2.HUMIDITY |  |  | -0.036 (-1.24) | -0.045 (-1.55) | -0.047 (-1.63) | -0.049+ (-1.74) | -0.054+ (-1.93) | -0.054+ (-1.96) | -0.050+ (-1.85) |
| L3.HUMIDITY |  |  |  | 0.007 (0.29) | 0.005 (0.21) | -0.005 (-0.20) | -0.010 (-0.42) | -0.016 (-0.66) | -0.012 (-0.51) |
| L4.HUMIDITY |  |  |  |  | 0.025 (1.03) | 0.018 (0.76) | 0.008 (0.36) | 0.005 (0.23) | 0.001 (0.04) |
| L5.HUMIDITY |  |  |  |  |  | 0.004 (0.19) | -0.005 (-0.22) | -0.016 (-0.76) | -0.021 (-1.00) |
| L6.HUMIDITY |  |  |  |  |  |  | 0.039+ (1.68) | 0.038 (1.56) | 0.032 (1.39) |
| L7.HUMIDITY |  |  |  |  |  |  |  | 0.024 (1.04) | 0.033 (1.37) |
| L8.HUMIDITY |  |  |  |  |  |  |  |  | 0.032 (1.46) |
| L0.TEMP_MAX | -0.000 (-0.10) | -0.000 (-0.03) | -0.000 (-0.08) | 0.000 (0.01) | 0.000 (0.07) | 0.000 (0.19) | 0.000 (0.09) | 0.000 (0.10) | 0.000 (0.34) |
| L1.TEMP_MAX |  | 0.000 (0.09) | 0.000 (0.18) | 0.000 (0.09) | 0.000 (0.01) | 0.000 (0.11) | 0.001 (0.70) | 0.001 (0.94) | 0.001 (1.14) |
| L2.TEMP_MAX |  |  | -0.002+ (-1.96) | -0.002+ (-1.84) | -0.002+ (-1.83) | -0.002 (-1.63) | -0.002 (-1.24) | -0.002 (-1.65) | -0.002+ (-1.76) |
| L3.TEMP_MAX |  |  |  | 0.001 (1.25) | 0.001 (1.30) | 0.001 (1.26) | 0.001 (1.03) | 0.001 (0.95) | 0.001 (0.56) |
| L4.TEMP_MAX |  |  |  |  | -0.001 (-0.58) | -0.000 (-0.21) | -0.001 (-0.69) | -0.000 (-0.43) | -0.000 (-0.38) |
| L5.TEMP_MAX |  |  |  |  |  | -0.002+ (-1.71) | -0.001 (-1.54) | -0.001 (-1.42) | -0.001 (-1.60) |
| L6.TEMP_MAX |  |  |  |  |  |  | -0.000 (-0.11) | 0.000 (0.42) | 0.000 (0.20) |
| L7.TEMP_MAX |  |  |  |  |  |  |  | -0.001 (-0.72) | -0.001 (-0.53) |
| L8.TEMP_MAX |  |  |  |  |  |  |  |  | 0.001 (0.64) |
| L0.TEMP_MIN | 0.000 (0.34) | 0.000 (0.21) | 0.000 (0.25) | 0.000 (0.11) | 0.000 (0.08) | -0.000 (-0.08) | 0.000 (0.14) | 0.000 (0.42) | -0.000 (-0.16) |
| L1.TEMP_MIN |  | 0.000 (0.24) | -0.000 (-0.12) | -0.000 (-0.24) | -0.000 (-0.26) | 0.000 (0.01) | -0.000 (-0.44) | -0.001 (-0.79) | -0.001 (-0.72) |
| L2.TEMP_MIN |  |  | 0.005*** (4.31) | 0.005*** (4.21) | 0.005*** (4.20) | 0.005*** (3.75) | 0.004** (3.20) | 0.004*** (3.49) | 0.004*** (3.41) |
| L3.TEMP_MIN |  |  |  | 0.000 (0.28) | 0.000 (0.29) | 0.000 (0.33) | 0.001 (0.99) | 0.001 (1.07) | 0.002 (1.28) |
| L4.TEMP_MIN |  |  |  |  | 0.001 (1.43) | 0.001 (1.15) | 0.001 (0.72) | 0.000 (0.38) | 0.000 (0.28) |
| L5.TEMP_MIN |  |  |  |  |  | 0.003* (2.27) | 0.002* (2.42) | 0.002* (2.09) | 0.002+ (1.72) |
| L6.TEMP_MIN |  |  |  |  |  |  | 0.001 (0.53) | 0.000 (0.31) | -0.000 (-0.10) |
| L7.TEMP_MIN |  |  |  |  |  |  |  | 0.001 (1.21) | 0.001 (0.67) |
| L8.TEMP_MIN |  |  |  |  |  |  |  |  | 0.001 (1.42) |
| Constant | 0.240*** (3.65) | 0.192+ (1.93) | 0.234+ (1.90) | 0.248+ (1.66) | 0.282+ (1.67) | 0.328* (2.04) | 0.196 (1.13) | 0.233 (1.43) | 0.230 (1.39) |
| Number of Observations | 6,588 | 6,588 | 6,588 | 6,586 | 6,568 | 6,524 | 6,468 | 6,381 | 6,223 |
| Number of States | 152 | 152 | 152 | 152 | 152 | 152 | 152 | 152 | 152 |
| Adjusted R-squared | 0.118 | 0.119 | 0.124 | 0.128 | 0.132 | 0.137 | 0.141 | 0.143 | 0.146 |

Note: +: $p < 0.10$ , \*: $p < 0.05$ , \*\*: $p < 0.01$ . t-statistics based on robust standard errors in parentheses. L0.UVI stands for the effect of UVI at time $t$ on the cumulative number of COVID-19 deaths at the same time, whereas L1.UVI, L2.UVI and L3.UVI (etc.) stand for the effect of UVI lagged by one, two, or three (etc.) weeks respectively. The same is true for the other variables.

**Table S8:** Number of Observations (Obs.) of Countries Used in Analysis

| Country | Obs. | Country | Obs. | Country | Obs. | Country | Obs. |
| --- | --- | --- | --- | --- | --- | --- | --- |
| Afghanistan | 47 | Cuba | 39 | Kazakhstan | 38 | Qatar | 41 |
| Albania | 42 | Cyprus | 42 | Kenya | 38 | Romania | 47 |
| Algeria | 55 | Czechia | 47 | Korea, South | 73 | Russia | 50 |
| Andorra | 47 | Denmark | 53 | Kosovo | 25 | San Marino | 53 |
| Angola | 31 | Djibouti | 28 | Kuwait | 34 | Sao Tome and Principe | 7 |
| Antigua and Barbuda | 31 | Dominican Republic | 50 | Kyrgyzstan | 33 | Saudi Arabia | 45 |
| Argentina | 48 | Ecuador | 50 | Latvia | 35 | Senegal | 37 |
| Armenia | 43 | Egypt | 61 | Lebanon | 59 | Serbia | 45 |
| Australia | 68 | El Salvador | 32 | Liberia | 34 | Sierra Leone | 15 |
| Austria | 55 | Equatorial Guinea | 16 | Libya | 27 | Singapore | 48 |
| Azerbaijan | 50 | Estonia | 44 | Liechtenstein | 34 | Slovakia | 37 |
| Bahamas | 35 | Eswatini | 22 | Lithuania | 48 | Slovenia | 46 |
| Bahrain | 53 | Ethiopia | 33 | Luxembourg | 51 | Somalia | 30 |
| Bangladesh | 43 | Finland | 48 | Malawi | 18 | South Africa | 42 |
| Barbados | 33 | France | 73 | Malaysia | 52 | Spain | 66 |
| Belarus | 38 | Gabon | 37 | Maldives | 9 | Sri Lanka | 41 |
| Belgium | 58 | Georgia | 34 | Mali | 26 | Sudan | 38 |
| Benin | 32 | Germany | 60 | Malta | 30 | Sweden | 58 |
| Bolivia | 40 | Ghana | 37 | Mauritius | 33 | Switzerland | 55 |
| Bosnia and Herzegovina | 46 | Greece | 54 | Mexico | 50 | Syria | 29 |
| Botswana | 21 | Guatemala | 37 | Moldova | 43 | Taiwan* | 73 |
| Brazil | 52 | Guinea | 23 | Monaco | 40 | Tanzania | 35 |
| Brunei | 41 | Guinea-Bissau | 12 | Montenegro | 34 | Thailand | 68 |
| Bulgaria | 43 | Guyana | 39 | Morocco | 49 | Togo | 42 |
| Burkina Faso | 41 | Haiti | 31 | Netherlands | 53 | Trinidad and Tobago | 37 |
| Burma | 24 | Honduras | 40 | New Zealand | 40 | Tunisia | 47 |
| Cabo Verde | 31 | Hungary | 47 | Niger | 31 | Turkey | 40 |
| Cameroon | 44 | Iceland | 50 | Nigeria | 46 | US | 69 |
| Canada | 60 | India | 58 | North Macedonia | 47 | Ukraine | 48 |
| Chad | 10 | Indonesia | 49 | Norway | 54 | United Arab Emirates | 49 |
| Chile | 47 | Iran | 61 | Oman | 38 | United Kingdom | 63 |
| China | 73 | Iraq | 56 | Pakistan | 50 | Uruguay | 37 |
| Colombia | 45 | Ireland | 51 | Panama | 41 | Uzbekistan | 36 |
| Congo (Brazzaville) | 36 | Israel | 48 | Paraguay | 43 | Venezuela | 37 |
| Congo (Kinshasa) | 40 | Italy | 73 | Peru | 45 | West Bank and Gaza | 43 |
| Costa Rica | 45 | Jamaica | 40 | Philippines | 73 | Yemen | 8 |
| Cote d'Ivoire | 40 | Japan | 73 | Poland | 47 | Zambia | 33 |
| Croatia | 50 | Jordan | 42 | Portugal | 49 | Zimbabwe | 31 |
| Total Number of Observations |  |  |  | 6,524 |  |  |  |

